# Early maternal plasma protein signatures identify preeclampsia risk in twin pregnancies

**DOI:** 10.64898/2026.09.28.26364186

**Authors:** Jiaxin Li, Zhuofan Fang, Wenmin Tian, Tianchen Wu, Yu Feng, Qi Wen, Xiaona Li, Jia Hao, Jun Ma, Yangyu Zhao, Yang Chen, Yuan Wei, Jing Hang

## Abstract

**BACKGROUND:** Twin pregnancies have a two- to three-fold higher risk of preeclampsia (PE), but early prediction remains poor because current biomarkers and prediction models are largely derived from singleton pregnancies. Defining the biological changes that precede PE in twins may facilitate the development of more effective risk assessment approaches.

**OBJECTIVE:** We aimed to characterize maternal plasma protein changes preceding PE in twin pregnancies and to explore whether early circulating proteins could serve as improved risk stratification for twin PE.

**STUDY DESIGN:** We performed a nested case-control study within a prospective twin pregnancy cohort at Peking University Third Hospital. A total of 84 women with twin pregnancies were included, comprising 40 pregnancies complicated by PE and 44 normotensive controls. Longitudinal maternal plasma samples were collected in the first, second, and third trimesters. In the discovery cohort, maternal plasma proteins were profiled by mass spectrometry to identify trimester-specific protein changes associated with PE. Candidate proteins were then validated in an independent validation set. We further assessed the functional relevance of selected proteins in trophoblast and endothelial cell models and constructed prediction models integrating biomarkers with maternal clinical characteristics.

**RESULTS:** Twin pregnancies complicated by PE showed distinct trimester-dependent protein alterations, with first- and second-trimester changes mainly involving immune regulation and glycolipid metabolism, and later changes enriched in cardiovascular-related pathways. Several proteins demonstrated early discriminatory patterns, including increased annexin A3, chitotriosidase-1, and ferritin light chain (FTL), together with decreased annexin A2 in the first trimester, whereas the conventional singleton markers (sFlt-1 and PAPP-A) did not differ. ELISA validation of these candidate biomarkers yielded an AUC of 0.93 for twin preeclampsia prediction; integration with standard maternal clinical factors further improved discriminatory performance to an AUC of 0.97. Accordingly, a multivariable model combining annexin A2, annexin A3, chitotriosidase-1, FTL, and maternal clinical factors achieved strong internal performance for early discrimination of PE in twin pregnancies (AUC, 0.97; 95% CI, 0.93–1.00). Moreover, annexin A2 knockdown impaired trophoblast migration and invasion and reduced inflammatory cytokine secretion in endothelial cells, supporting a role in placental development and maternal vascular adaptation.

**CONCLUSION:** Twin pregnancies that later develop PE exhibit early and progressive maternal plasma protein alterations that can be detected before clinical disease onset. These findings identify candidate biomarkers and biologically relevant pathways that may help explain why PE in twins differs from singleton disease and may support future twin-specific risk stratification strategies. These results require external validation before clinical application.

## Introduction

Preeclampsia (PE) is a placenta-mediated pregnancy complication syndrome characterized by new-onset hypertension and multisystem dysfunction, and remains a leading cause of elevated maternal and perinatal morbidity and mortality.^1, 2^ This issue is particularly relevant in twin pregnancies, where the risk of PE is approximately two- to threefold higher than in singleton pregnancies, with reported incidences ranging from 13% to 37%. For obstetricians and maternal-fetal medicine specialists, PE in twin pregnancies presents a practical clinical challenge. Most currently available tools for PE risk assessment and prevention were developed and validated primarily in singleton pregnancies, and their performance is reduced when applied directly to twin gestations.^3^ For instance, a competing-risks model based on maternal characteristics and medical history exhibits substantially decreased performance in twins, while the widely used sFlt-1/PLGF ratio shows limited predictive utility in this population.^4, 5^ In addition, many biomarker-based assessments are used after clinical risk has become apparent, whereas guidelines recommend initiating oral aspirin prophylaxis in high-risk women before 16 weeks of gestation.^3, 6, 7^ These limitations underscore the critical need to elucidate the molecular pathogenesis underlying twin PE and to develop twin-specific prediction strategies for more accurate diagnosis and improved clinical management.

Twin PE displays distinct pathophysiological features, including excessive placental mass,^8, 9^ increased maternal inflammatory activation,^10, 11^ and 40%–50% higher hemodynamic burden relative to singleton PE.^12–14^ These changes may impair maternal cardiovascular adaptation and contribute to disease development. Furthermore, twin PE is also characterized by profound metabolic disorders, as reflected by markedly increased maternal lipid levels.^15^ Meanwhile, the role of gestational diabetes mellitus (GDM) as a risk factor for twin PE remains controversial. Severe cases of twin PE are associated with a higher prevalence of GDM,^16^ while other evidence indicates that GDM exerts a relatively smaller impact on adverse pregnancy outcomes in twins than in singletons.^17^ These differences indicate that PE in twins may involve distinct maternal-placental adaptive responses and metabolic disruption, rather than a simple amplification of the mechanisms observed in singleton PE.

Given the biological complexity of twin PE, maternal plasma proteomics provides a useful approach to characterize the disease-related molecular changes. High-throughput proteomic studies have established that maternal plasma proteins change substantially across normal pregnancy and can be used to model gestational biology.^18, 19^ Previous studies have also identified dysregulated proteins involved in angiogenesis, inflammation, and coagulation,^20–22^ and have revealed candidate early biomarkers for PE, such as matrix metalloproteinase-7 (MMP7) and glycoprotein IIb/IIIa complex.^23^ In addition, maternal characteristics and circulating biomarkers can be used to predict PE across gestation.^24^ Although these studies provide an important clinical and biological foundation of PE, most have focused on singleton pregnancies. Longitudinal maternal plasma protein changes specific to twin pregnancies that later develop PE remain insufficiently defined.

In this study, we performed a trimester-resolved proteomic profiling of maternal plasma samples collected during the first, second, and third trimesters from a cohort of 84 twin pregnancies. Comparative analyses between normotensive twin pregnancy and twin PE led to the identification of differential protein expression patterns in each trimester and their associated biological functions. We discovered that maternal plasma protein alterations can precede the clinical onset of PE in twin pregnancies, and identified potential early biomarkers for distinguishing twin pregnancies at elevated PE risk. By focusing specifically on twin pregnancies, these results provide a molecular basis for early risk stratification in this high-risk obstetric population.

## Materials and Methods

### Study population and sample collection

This nested case-control study was conducted within a prospective twin pregnancy cohort established at Peking University Third Hospital. Women aged 18–45 years with twin pregnancies (<14 weeks of gestation) were recruited between 2023 and 2024. Preeclampsia (PE) was defined according to the criteria of the International Society for the Study of Hypertension in Pregnancy. For proteomic and validation study, we enrolled a total of 40 twin PE cases and 44 matched normotensive controls based on maternal age, pre-pregnancy BMI, parity, conception method, and chorionicity. Pregnancies complicated by selective fetal growth restriction, twin-twin transfusion syndrome, twin reversed arterial perfusion sequence, or twin anemia-polycythemia sequence were excluded. Maternal blood samples were collected from each participant at predetermined time points in the first, second, and third trimesters alongside routine prenatal assessments. Placental tissues were obtained at delivery. All samples were processed using standardized protocols and stored at −80°C until analysis. Laboratory analyses were conducted with investigators blinded to clinical outcomes.

### Proteomic profiling of maternal plasma

Plasma proteomic profiling was performed using data-independent acquisition-based liquid chromatography-tandem mass spectrometry (DIA LC-MS/MS). High-abundance proteins were depleted prior to analysis to enhance detection of low-abundance proteins. Protein identification and quantification were conducted using Spectronaut software, with a false discovery rate of <1% at both peptide and protein levels. The whole study procedure of this work is summarized in Figure 1.

**Figure 1.** Study design and longitudinal plasma proteome profiling of twin pregnancy. Maternal plasma samples from each gestational trimesters of twin pregnancies complicated by preeclampsia (PE, n = 26) and normotensive controls (N, n = 34) were subjected to low-abundance proteomic analysis using data-independent acquisition liquid chromatography–tandem mass spectrometry (DIA LC-MS/MS). Pathway enrichment and multiple analytical strategies were applied to identify candidate biomarkers, which were further verified using ELISA in an independent cohort (14 PE vs 10 N). Further investigation was completed by cellular and histochemical assays. Predictive model tailored for twin PE was subsequently constructed. T1, T2, and T3 indicate the first, second, and third trimesters.

### Bioinformatics analysis

Differentially expressed proteins were identified using |fold change| ≥ 2.0 and *p*-value < 0.05. Principal component analysis and Pearson’s correlation coefficient were applied to evaluate sample reproducibility. Statistical significance (*p* < 0.05) was determined using Student’s *t*-test. The *p*-values were adjusted using the Benjamini-Hochberg multiple testing correction. All statistical analyses were performed using the t-test_ind function from the SciPy library in Python (version 3.7.6). KEGG database was utilized for protein annotation of differentially expressed proteins, and pathway enrichment analysis was performed using a two-sided Fisher’s exact test conducted with the fisher_exact function from scipy.stats in Python (version 3.7.6). KEGG pathways with an adjusted *p*-value < 0.05 were considered statistically significant.

### Enzyme-linked immunosorbent assay (ELISA)

Circulating protein levels for annexin family members, chitotriosidase-1, ferritin light chain, and S100 calcium-binding protein A12 (S100-A12) in maternal plasma from PE cases and controls were quantified to validate the proteomic results. In addition, cytokine secretion (including IL-6 and IL-8) was measured in HUVEC cell supernatants to evaluate the downstream inflammatory effects of annexin A2.

### Cell culture and transfection

Human trophoblast (HTR-8/SVneo) and endothelial (HUVEC) cell models were used to investigate the functional role of annexin A2. Cells were maintained under standard culture conditions. Annexin A2 expression was modulated by siRNA-mediated knockdown or plasmid-based overexpression, followed by downstream analyses of trophoblast function and endothelial inflammatory responses.

### Real-time quantitative PCR (RT-qPCR)

Total RNA was extracted from HTR-8/SVneo cells and reverse-transcribed into cDNA. RT-qPCR was performed using SYBR Green chemistry following standard protocols. Relative mRNA expression levels were calculated using the 2^^−ΔΔCt^ method and normalized to GAPDH.

### Wound-healing and transwell assays

The migratory and invasive capacities of HTR8/SVneo trophoblast cells were evaluated following annexin A2 knockdown or overexpression. Cell migration was assessed using wound-healing assays, and invasion was examined using transwell assays. Quantitative analyses were performed based on wound closure and the number of invading cells, respectively.

### Western blotting

Protein expression levels in HTR-8/SVneo cells were assessed by Western blot analysis. Annexin A2 expression was detected and normalized to GAPDH as a loading control. Protein bands were visualized using chemiluminescence and quantified by densitometric analysis. Uncropped immunoblots are provided in the Supplemental Figure 10.

### Immunohistochemistry staining

Twin placental tissue samples obtained from four normal and four PE twin pregnancies were collected exclusively for immunohistochemistry validation. Paraffin-embedded placental sections were processed and stained for annexin A2, annexin A3, chitotriosidase-1, and ferritin light chain. Immunoreactive signals were visualized and semi-quantitatively analyzed based on representative microscopic fields.

### Malondialdehyde analysis

Malondialdehyde levels were measured in homogenized placental samples from twin pregnancies. Malondialdehyde concentrations were determined following standard commercial kit instructions.

### Statistical analyses

Continuous variables were presented and compared using parametric or non-parametric methods according to normality, with adjusted post hoc pairwise comparisons for multi-group analyses. Categorical variables were summarized as frequencies with percentages and compared using the chi-square test or Fisher’s exact test. All statistical tests were two-sided, with P < 0.05 considered statistically significant.

### Construction of prediction model

Candidate biomarkers were initially selected from the training set (26 PE vs. 34 controls) based on distinct gestational expression patterns in the proteomic profile and relevance to PE pathogenesis. Six machine learning algorithms, including logistic regression, decision tree, naive Bayes, support vector machine (SVM), XGBoost, and k nearest neighbors (KNN), were used to evaluate the predictive performance of the candidate biomarkers. After ELISA validation, the chosen biomarkers were used for prediction models construction with four configurations: univariable biomarker models, multivariable biomarker models, clinical risk factor models, and comprehensive models combining biomarkers with clinical risk factors (maternal age, pre-pregnancy BMI, parity, conception mode, and mean arterial pressure). Model performance was assessed via receiver operating characteristic (ROC) analysis with area under the curve (AUC), and optimal thresholds were determined by the Youden’s index.

### Supplemental Materials and Methods

Detailed experimental procedures, including sample preparation, LC-MS/MS parameters, molecular assays, and cell-based experiments, are provided in the Supplemental Data.

## Results

### Clinical Characteristics and Longitudinal Maternal Plasma Protein Profiling

A total of 40 twin pregnancies complicated by PE and 44 matched normotensive controls (N) were included to profile the plasma proteome dynamics across twin gestations and to identify PE-associated alterations. Of those, 26 PE cases and 34 N participants comprised the discovery cohort for proteomic analysis, while 14 PE cases and 10 N participants were assigned to the validation cohort. Baseline characteristics were comparable between groups (Table 1). Mean arterial pressure (MAP) at 11–14 weeks was elevated in the PE group (*p* = 0.002), and the incidence of GDM and the proportion of first-delivered fetuses with birth weight below the 10^th^ percentile was slightly higher (*p* < 0.001), suggesting the presence of early metabolic and vascular alterations before clinical disease onset.

**Table 1.**
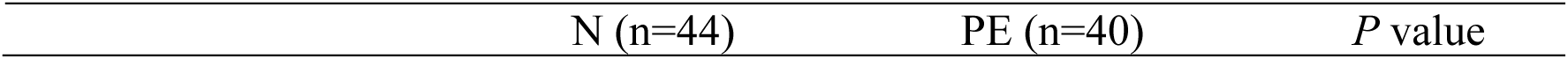
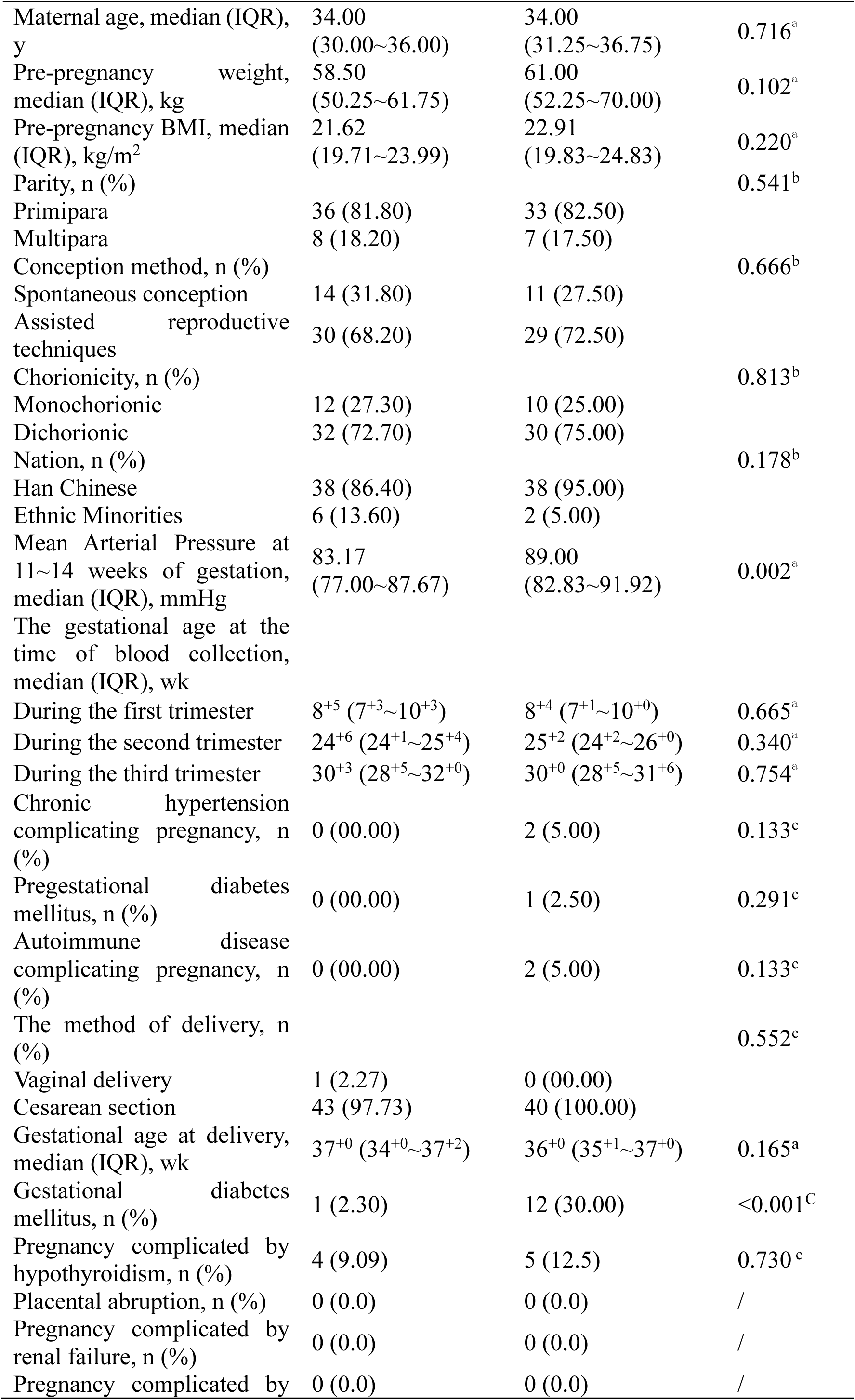
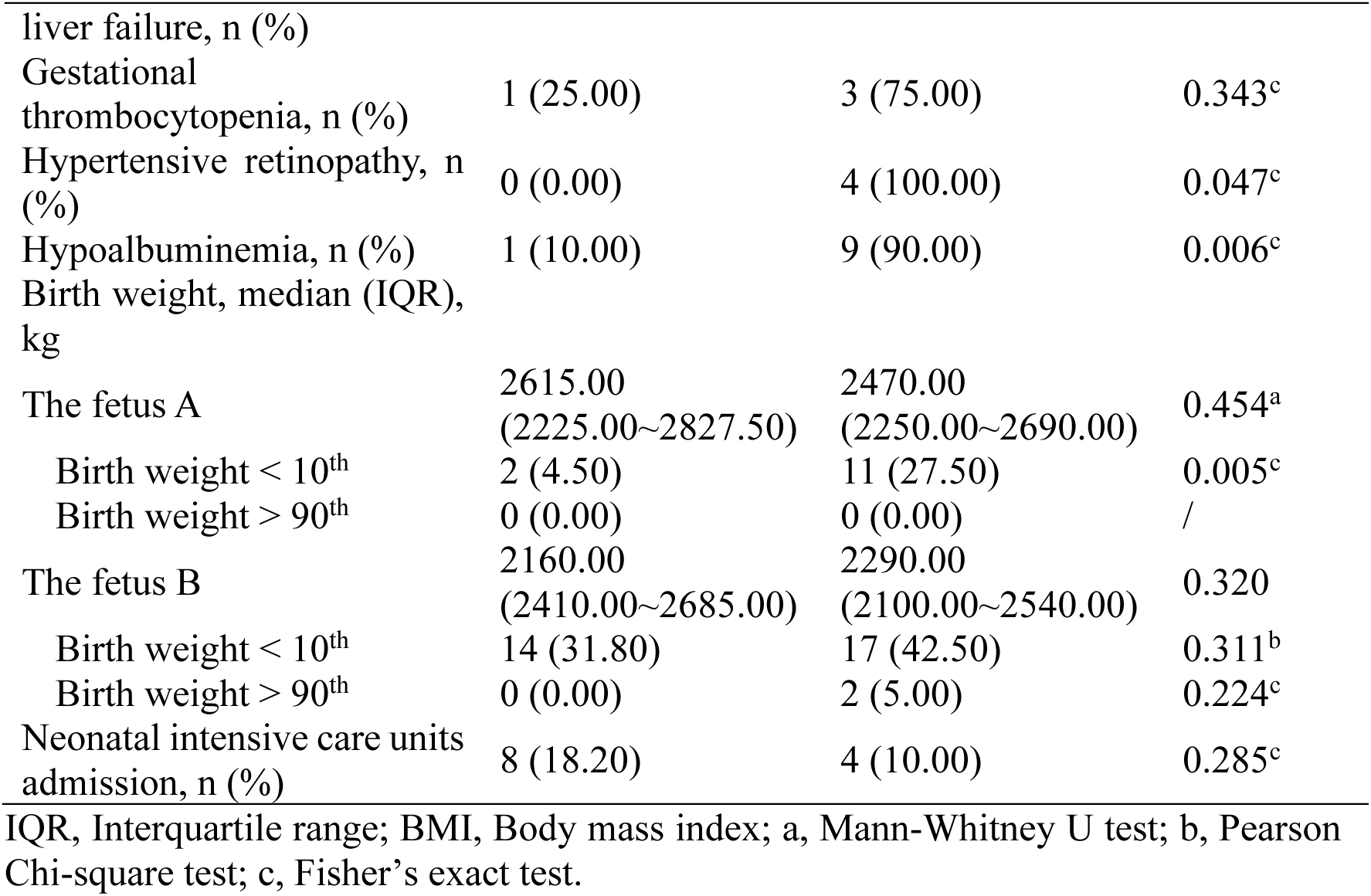
Clinical characteristics of twin pregnancies with and without preeclampsia.

Proteomic profiling quantified 1,486 maternal plasma proteins across gestation (Supplemental Figure 1A). Principal component analysis of the normalized proteomic dataset demonstrated a clear separation between PE and N groups at each gestational stage, reflecting global proteomic shifts (Supplemental Figure 1B). Pairwise comparison between PE and N at each trimester identified 71 upregulated and 67 downregulated proteins in the first trimester, 67 upregulated and 62 downregulated proteins in the second trimester, and 48 upregulated and 38 downregulated proteins in the third trimester, respectively (Supplemental Figure 1C–E), indicating the greatest divergence occurring during early and mid-gestation.

### Trimester-Specific Protein Changes Distinguish Twin Pregnancies that Develop Preeclampsia

Among normotensive twin pregnancies, protein changes between the first and second trimesters were primarily enriched in cholesterol metabolism, inflammatory responses, and ferroptosis (Supplemental Figure 2A), consistent with their role in maternal-fetal immune tolerance.^25^ Additionally, proteins altered between the third trimester and earlier stages were mapped predominantly to metabolic and cardiovascular pathology pathways (Supplemental Figure 2, B and C). These analyses provide a comprehensive resource for understanding the molecular processes underlying physiological adaptations in normotensive twin pregnancy, establishing a baseline against which pathological deviations can be contrasted.

In twin pregnancies that subsequently developed PE, proteomic alterations became progressively more pronounced across gestation. Comparisons between first and second trimester samples (Supplemental Figure 3A), and between first and third trimester samples (Supplemental Figure 3B), showed enrichment mainly in lipid and glucose metabolism, complement activation, and cytokine signaling related pathways. Furthermore, differences between mid- and late trimester exhibited continued divergence in protein expression (Supplemental Figure 3C). These proteomic patterns suggest that twin PE is accompanied by early immune and metabolic dysregulation followed by increasing cardiovascular and hemodynamic stress.

Direct comparison of PE and normotensive twin pregnancies showed that in the first (PE1 vs. N1) and second trimesters (PE2 vs. N2), differences in twin PE were significantly enriched in pathways related to glucose metabolism, including glycolysis/gluconeogenesis, carbon metabolism, and the tricarboxylic acid cycle (Figure 2A, B). By the third trimester (PE3 vs. N3), differentially expressed proteins shifted to pathways predominantly linked to viral myocarditis and type I diabetes mellitus (Figure 2C), suggesting the convergence of late-gestation PE with chronic metabolic and cardiovascular disorders.

**Figure 2.** Differentially expressed proteins and the associated pathway enrichment between twin PE and normotensive twin pregnancies across three trimesters. (A– C) Heatmaps and chord diagrams demonstrating DEPs and enriched KEGG pathways between PE1 vs. N1 (A), PE2 vs. N2 (B), and PE3 vs. N3 (C).

### Clinically Relevant Protein Trajectories in Twin Preeclampsia

Since PE usually becomes clinically apparent later in pregnancy, we focused on proteins whose expression diverged in the second and third trimesters. Proteins that increased later in PE were enriched in cholesterol metabolism, PPAR/TGF-beta/PI3K- Akt signaling, and platelet activation (Supplemental Figure 4A). Conversely, proteins that decreased later mapped mainly to the complement and coagulation cascade and metabolic homeostasis pathways (Supplemental Figure 4B). Representative proteins increasing in the second and third trimester included 6-phosphogluconolactonase^26^ (6PGL), a key pentose phosphate pathway enzyme crucial for glucose metabolism, and DCBLD2 (discoidin, CUB, and LCCL domain-containing protein 2),^27^ which regulates platelet activation and cell adhesion (Supplemental Figure 4C). HLA-A (Major Histocompatibility Complex, Class I, A) and IGLV7-46 (immunoglobulin lambda variable 7-46) increasing in the third trimester indicate a potential dysregulated cellular immunity (Supplemental Figure 4D). These findings link the plasma protein signal to processes that are clinically relevant to placental function, endothelial activation, and maternal cardiovascular adaptation.

### Discovery of Early Candidate Biomarkers and Predictive Model Development Specific for Twin PE

Currently, the increased levels of secreted fms-like tyrosine kinase-1 (sFlt-1), placental growth factor (PlGF), Endoglin (Eng), and leptin (LEP), along with the decreased levels of pregnancy-associated plasma protein A (PAPP-A), are widely used as conventional early predictive indicators for singleton PE.^28–30^ However, in our cohort, none of these markers showed significant expression differences between the twin PE and N groups in the first trimester (Supplemental Figure 5). This discrepancy raises the possibility that conventional biomarkers may be less applicable for early prediction of twin PE, pointing to the need for further exploration of twin-specific predictive approaches.

To identify early gestation proteomic alterations associated with twin PE, we performed feature selection based on trimester-specific differential expression analysis (Figure 3A–H). Both annexin A1 and annexin A3 were increased in the first trimester but declined thereafter in PE, contrasting with their progressive increase in normotensive twins (Figure 3A, B). Ferritin light chain (FTL) exhibited significant elevation in early stage of twin PE but no difference later (Figure 3C). Moreover, S100 calcium-binding protein A12 (S100-A12) and chitotriosidase-1 presented an inverted trajectory relative to normotensive controls (Figure 3D, E). Immunoglobulin kappa variable 1-16 (KV116), Albumin (ALB), and latent-transforming growth factor beta-binding protein 2 (LTBP2) also showed distinct longitudinal patterns (Figure 3F–H).

**Figure 3.** Machine learning-based early predictive model development specific for twin PE. (**A–H**) Representative proteins with distinct gestational dynamics. annexin A1 (ANXA1) (**A**); annexin A3 (ANXA3) (**B**); ferritin light chain (FTL) (**C**); S100 Calcium Binding Protein A12 (S100-A12) (**D**); chitotriosidase-1 (CHIT1) (**E**); immunoglobulin kappa variable 1-16 (KV116) (**F**); albumin (ALB) (**G**); latent-transforming growth factor beta-binding protein 2 (LTBP2) (**H**). The y-axis represents the normalized protein expression. Two-way ANOVA was used for statistical analysis. (**I**) An XGBoost machine learning-based early predictive model utilizes a total of the best-performing predictive proteins. Sens, sensitivity. Spec, specificity. * *P* < 0.05, ** *P* < 0.01, **** *P* < 0.0001.

To evaluate the predictive value of these differentially expressed proteins, we employed six machine learning algorithms, among which XGBoost demonstrated superior performance with the highest discriminative ability and robustness against overfitting (Supplemental Figure 6A–H, Supplemental Table 1). Individually, annexin A1, annexin A3, FTL, S100-A12, and chitotriosidase-1 showed promising discriminatory power (AUCs ranging from 0.63 to 0.91; Supplemental Figure 6A–E, Supplemental Table 1). The combined model integrating these five top-performing candidates biomarkers yielded even more exceptional predictive power, achieving an AUC of 0.96 (95% CI: 0.92–1.00) (Figure 3I), significantly outperforming each single marker. These results suggest the predictive potential of these candidate proteins that required further validation using XGBoost.

### Validation of Candidate Biomarkers and Improved Early Prediction Model

The plasma levels of the above-mentioned candidates were then validated by ELISA in the independent cohort (Figure 4A–E). Among them, only annexin A3, chitotriosidase- 1, and FTL exhibited consistent changes as detected by mass spectrometry (Figure 4B– D). Given the homology of annexin A2 to annexins A1/A3 and its distinctive expression trends in twin PE during our manual curation (Supplemental Figure 7A), we also examined the levels of annexin A2. Notably, it presented a more pronounced decrease in early twin PE (Figure 4F), suggesting its potential as a stronger candidate for predictive targeting.

**Figure 4.** Validation and predictive performance of early biomarkers for twin PE. (**A–E**) Protein expression level validation using ELISA of annexin A1 (ANXA1) (**A**), annexin A3 (ANXA3) (**B**), chitotriosidase-1 (**C**), ferritin light chain (FTL) (**D**), S100 Calcium Binding Protein A12 (S100-A12) (**E**), and annexin A2 (ANXA2) (**F**) in maternal plasma. Student’s *t*-test was used for statistical comparison in A–F. (**G–H**) Immunohistochemistry staining of placental villi from twin placental samples and the corresponding quantification of ANXA2 (**G**), ANXA3 (**H**), CHIT1 (**I**), and FTL (**J**). Scale bars = 50 μm. One-way ANOVA was used. Data represents four independent biological replicates. (**K–P**) Receiver operating characteristic (ROC) curves illustrating predictive performance of: ANXA2 (Sens = 0.89, Spec = 0.76, AUC = 0.88) (**K**), ANXA3 (Sens = 0.83, Spec = 0.78, AUC = 0.85) (**L**), CHIT1 (Sens = 0.84, Spec = 0.82, AUC = 0.86) (**M**), FTL (Sens = 0.84, Spec = 0.74, AUC = 0.84) (**N**), combined biomarkers (ANXA2+ANXA3+CHIT1+ FTL) (Sens = 1.00, Spec = 0.76, AUC = 0.93) (**O**), and biomarker panel with clinical parameters (Sens = 0.92, Spec = 0.94, AUC = 0.97) (**P**). Clinical parameters include maternal age, pre-pregnancy BMI, parity status, conception method (natural vs. ART), and mean arterial pressure (MAP = [2×diastolic plasma pressure + systolic plasma pressure]/3). Sens, sensitivity. Spec, specificity. * *P* < 0.05, ** *P* < 0.01, *** *P* < 0.001, **** *P* < 0.0001.

Immunohistochemistry on placental tissues collected at delivery supported the plasma findings (Supplemental Table 2). Annexin A2 was significantly reduced in the placentas of twin pregnancies complicated by PE (Figure 4G), along with the marked upregulation of annexin A3, chitotriosidase-1, and FTL (Figure 4H–J). These findings indicate the altered plasma levels may reflect placental dysfunction and therefore reinforce their biomarker potential for early twin PE prediction.

Building on the ELISA results, we refined the XGBoost predictive model. When evaluated individually, annexin A2 demonstrated superior predictive capability (AUC = 0.88, 95% CI: 0.78–0.98), surpassing annexin A3, chitotriosidase-1, and FTL (Figure 4K–N). Integration of these four biomarkers improved model performance, achieving an AUC of 0.93 (95% CI: 0.85–1.00; Figure 4O). To enhance predictive accuracy, we further incorporated key maternal clinical risk factors, including maternal age, pre-pregnancy BMI, parity, conception method, and first-trimester mean arterial pressure (MAP). Clinical risk factors alone yielded an AUC of 0.842 (95% CI: 0.743–0.940; Supplemental Figure 6I), and combining these factors with the proteomic biomarker panel improved predictive performance to an AUC of 0.97 (95% CI: 0.93–1.00; Figure 4P, Supplemental Table 3), suggesting that the combination of proteomic biomarkers and clinical variables may facilitate more accurate early risk stratification in twin pregnancies.

### Annexin A2 links the biomarker signal to impaired placental function

To elucidate the role of annexin A2 in twin PE pathogenesis, we first tested its longitudinal expression pattern. In normotensive twins, circulating annexin A2 was steadily increased, whereas this rise was significantly blunted in twin PE (Figures 4F, 5A). Given the central role of placental dysfunction in PE pathophysiology, we next investigated the functional relevance of annexin A2 in extravillous trophoblast (EVT) cells. Annexin A2 knockdown—confirmed at both mRNA and protein levels via reverse transcription quantitative real-time polymerase chain reaction (RT-qPCR) and western blot, respectively (Figure 5B–D)—significantly suppressed HTR-8/SVneo cell migration and invasion (Figure 5E–H). Conversely, annexin A2 overexpression enhanced EVT function (Supplemental Figure 7B-H), indicating the positive impact of annexin A2 in placental development.

**Figure 5.** Functional characterization of annexin A2 (ANXA2) in twin PE. (**A**) ELISA validation of ANXA2 in T2 and T3. (**B**) Knockdown of ANXA2 in HTR- 8/SVneo trophoblast cells confirmed by RT-qPCR. Data represents four independent biological replicates. Blank, untransfected; siNC, negative control siRNA; siANXA2- 1 and siANXA2-2, two ANXA2-specific siRNAs. (**C–D**) Western blot validation of ANXA2 knockdown (**C**), with semi-quantitative analysis based on densitometry (**D**). (**E–F**) Migration assay of HTR-8/SVneo cells following ANXA2 silencing (**E**) was quantitatively assessed (**F**) using a quantitative analysis based on scratch closure area. Data represents three independent biological replicates. Scale bars: 250 μm. (**G–H**) Invasion assay of HTR-8/SVneo cells after ANXA2 knockdown (**G**) through a quantitative analysis based on the number of invading cells (**H**). Data represents three independent biological replicates. Scale bars: 50 μm. (**I–L**) Impact of ANXA2 knockdown on cytokine release in human umbilical vein endothelial cells (HUVEC) cells. Interleukin-6 (IL-6) (**I**) and IL-8 (**L**) were tested. Data represents six independent biological replicates. * *P* < 0.05, ** *P* < 0.01, *** *P* < 0.001, **** *P* < 0.0001. T1, T2, and T3 indicate the first, second, and third trimesters.

Considering that annexin A2 is a multifunctional calcium-dependent phospholipid-binding protein involved in vascular remodeling and complement-mediated immune regulation,^31^ we further assessed its influence on inflammatory responses. Silencing of annexin A2 in human umbilical vein endothelial cells (HUVECs) reduced secretion of pro-inflammatory cytokines interleukin-6 (IL-6) and interleukin-8 (IL-8) (Figure 5I, J).

### Comparison between monochorionic and dichorionic twins

Chorionicity, categorized as either monochorionic (MC) or dichorionic (DC) twins, is another critical determinant of twin pregnancy outcomes. However, their incidence of PE and pathophysiological mechanisms remain inconclusive.^32, 33^ Our chorionicity- stratified enrichment analyses displayed that early proteomic profiles did not differ between MC and DC twins under normal conditions (Supplemental Figure 8A), but distinct differences emerged in the context of PE (Supplemental Figure 8A and B). We therefore analyzed PE cases separately by chorionicity type. In MC pregnancies with PE (MC-PE), the ferroptosis pathway was specifically enriched, accompanied by dysregulation of key proteins involved in iron homeostasis, such as FTL and transferrin receptor (TFRC) (Supplemental Figure 8C–E).^34^ Conversely, these alterations were not observed in DC-PE across gestation (Supplemental Figure 8F–H). These findings implicate a potential role of ferroptosis in the pathogenesis of MC-PE.

### Comment

#### Principal findings

This longitudinal study demonstrates that among women with twin pregnancies, those who subsequently developed preeclampsia (PE) demonstrated distinct maternal plasma protein changes that were detectable before clinical disease onset and became progressively more pronounced throughout gestation. The early-trimester signal of several proteins, including annexin A2, annexin A3, chitotriosidase-1, and ferritin light chain (FTL), distinguished pregnancies that later developed PE from normotensive twin pregnancies. These markers improved internal risk discrimination when combined with routine maternal clinical factors. Functional experiments further supported the biological relevance of annexin A2 in trophoblast migration and invasion. Together, these findings suggest that the pathophysiologic processes leading to PE in twin pregnancies begin well before clinical diagnosis and may be applied as maternal blood-based biomarkers.

#### Results in the context of what is known

Twin pregnancies carry a substantially higher risk of hypertensive disorders of pregnancy, particularly PE, than singleton pregnancies. However, most currently available screening algorithms, risk models, and biomarker-based prediction strategies were developed and validated in singleton pregnancies. Their reduced performance in twin gestations creates a clinically important unmet need for twin-specific prediction approaches. In this context, our study provides an innovative longitudinal plasma proteomic analysis focused on twin PE and identifies distinguished molecular features that may support earlier risk stratification in this high-risk population.

Previous studies have demonstrated dynamic changes in maternal plasma proteins throughout normal pregnancy, supporting their value for characterizing gestational biology.^18, 19^ Other studies have also shown that maternal clinical characteristics together with circulating biomarkers can predict PE at different stages of pregnancy.^24^ The present study extends this evidence to twin pregnancies through maternal-plasma proteomics with a longitudinal, trimester-resolved design and ELISA validation. In addition, tissue localization and functional experiments support the biological relevance of selected markers, particularly annexin A2. These findings provide twin-specific evidence that clinically relevant protein alterations can precede the onset of PE.

#### Mechanistic insights relevant to obstetric practice

Several proteins identified in this study showed distinct gestational patterns before the onset of overt PE. This is clinically important because PE prevention and surveillance strategies are most useful when risk can be recognized before maternal hypertension becomes apparent. In twin pregnancies, interpretation of circulating biomarkers is complicated by larger placental mass and by twin-specific placental complications. To minimize these confounding possibilities, our cohort excluded all pregnancies complicated by selective fetal growth restriction, twin-twin transfusion syndrome, twin reversed arterial perfusion sequence, and twin anemia-polycythemia sequence. Therefore, the observed proteomic alterations are more likely to represent disease-specific molecular changes rather than nonspecific influences of twin gestation or other placental disorders.

Among these potential biomarkers, the glucocorticoid-regulated annexins were prominent. The elevated annexin A1 in singleton PE^35^ inhibits trophoblast inflammation^36^ and the upregulation of annexin A3 is associated with hypertension.^37^ ^38^ Annexin A2 is abundant at the maternal-fetal interface and crucial for trophoblast decidualization,^39^ but reports in singleton PE are conflicting: some observed increased annexin A2 in acute PE placenta,^40^ while others described decreases linked to placental thrombosis^41, 42^ and reduced maternal plasma levels in late PE.^43^ Our functional assays showed that annexin A2 knockdown suppressed trophoblasts invasion and migration and decreased endothelial pro-inflammatory cytokine secretion (Figure 5E–J), supporting its functional relevance in maternal-fetal immune tolerance during proper placentation. These findings underscore annexin A2 as a context-dependent regulator and support its dual value as a biomarker and therapeutic target in twin PE.

We also identified FTL as a novel early biomarker with strong predictive potential. FTL, an essential component for iron storage and oxidative homeostasis,^44^ is downregulated in both the maternal blood and placental villi of singleton PE pregnancies.^45^ In contrast, we observed an increased maternal plasma FTL levels in early twin PE (Figure 3C), and its localization was predominant to placental villi but not decidua (Figure 4J; Supplemental Figure 9A). We performed malondialdehyde assays to assess placental oxidative status and observed significantly higher levels in the placentas of twin pregnancies with PE (Supplemental Figure 9B, indicating enhanced oxidative stress. The discrepancy between FTL levels and malondialdehyde trends suggest that although lipid peroxidation is elevated in twin PE, the concurrent increase in FTL levels in both placentas and maternal plasma may reflect a compensatory maternal adaption to counteract oxidative damage. These divergent findings underscore mechanistic dissimilarities between twin and singleton PE and further support the potential utility of FTL as a twin PE-specific biomarker.

Beyond their biological relevance, these proteins may also be clinically important. Their identification supports the concept that biologically meaningful alterations precede the clinical manifestations of twin PE. If validated in independent cohorts, such biomarkers may contribute to future approaches for early risk assessment and stratification in this high-risk population.

#### Prediction model, overfitting, and the need for external validation

From a methodological perspective, comparative analysis of multiple machine learning algorithms revealed that XGBoost achieved the best performance in this cohort (Supplemental Table 1). Through internal validation, we established a preliminary early prediction model for twin PE. Unlike most currently available PE prediction tools, which were developed in singleton cohorts, our model was derived entirely from twin gestations and therefore may better capture the unique biological features underlying PE in this population. Notably, annexin A2 alone demonstrated satisfactory predictive efficacy. Subsequent incorporation of annexin A3, chitotriosidase-1, and FTL further improved performance. The addition of maternal risk factors yielded an AUC of 0.97, exemplifying the power of combining proteomic biomarkers with clinical parameters for obstetric risk prediction.^29^

This apparent predictive performance should be interpreted with caution. Proteomic studies of twin pregnancies usually include modest sample sizes, and the number of PE cases is often limited. When many circulating proteins are evaluated in a relatively small cohort, a machine learning model may identify patterns that fit the study population very well but do not perform equally well in other clinical settings. Internal validation provides an initial estimate of model stability, Therefore, we further examined candidate proteins from the discovery phase in an independent ELISA cohort, and the biomarker signal remained encouraging.

However, both the biomarker identification and predictive model establishment should be considered hypothesis-generating and none of them are ready for clinical implementation at this stage. External validation in independent, larger, multicenter, and ethnically diverse twin populations, with prospective sampling before diagnosis, is required before incorporation into routine obstetrical practice. Future validation of these biomarkers may facilitate more individualized surveillance approaches and improve patient counseling through more accurate estimation of disease risk.

#### Clinical implications

Twin pregnancies are associated with a substantially higher risk of PE compared with singleton gestations, yet traditional prediction models developed for singletons demonstrate limited performance in this high-risk population. This study implies that PE risk assessment in twin pregnancies may require twin-specific biomarkers and prediction models. Twin gestations differ from singleton gestations in placental mass, maternal cardiovascular load, inflammatory activation, and metabolic adaptation. These differences may partly explain why prediction models developed in singleton pregnancies often show reduced performance when applied to twins. In this study, annexin A2, annexin A3, chitotriosidase-1, and FTL were measurable in maternal plasma and showed early alterations before the clinical onset of PE. A multivariable prediction model integrating these four proteomic biomarkers with maternal clinical characteristics achieved encouraging predictive performance in this study. These findings support the concept that biologically meaningful alterations occur before the clinical manifestations of twin PE and that these alterations can be detected in maternal plasma.

If validated in independent cohorts, a maternal blood-based panel incorporating these candidates may contribute to earlier risk stratification in twin pregnancies. Such a tool could help clinicians identify women who may benefit from closer blood pressure monitoring or more individualized counseling regarding PE risk. It may also help refine surveillance strategies in twin pregnancies. However, the present findings remain exploratory and further prospective validation is warranted potential clinical translation.

#### Research implications

This work establishes a comprehensive proteomic reference for healthy and PE twin pregnancies and highlights candidate pathways for mechanistic investigation, including immunity, ferroptosis, and cardiometabolic stress responses. Annexin A2 emerged as a particularly relevant candidate because it showed high predictive value and functional involvement in trophoblast behavior. FTL also warrants further study because its early increase in twin PE differs from previous patterns in singleton PE and may reflect twin-specific oxidative or metabolic adaptation. Future studies should evaluate the causality of these biomarkers or pathways, assess biomarker stability and predictive thresholds across diverse populations.

#### Strengths and limitations

A major strength of this study is its focus on twin pregnancies, a high-risk but understudied population in maternal plasma proteomic research. The longitudinal assessment of maternal plasma proteins across all three trimesters enabled characterization of disease-related molecular trajectories that cannot be captured by cross-sectional studies. Another strength is the systematic comparison of multiple machine learning algorithms and the integration of proteomic biomarkers with maternal clinical characteristics, which supported the development of a preliminary twin-specific prediction framework.

Limitations include the single-center cohort and modest sample size inherent to twin pregnancy studies, which may restrict generalizability. External validation in larger, diverse populations is warranted. While the mechanistic role of annexin A2 was explored, the causal contributions of other biomarkers require further investigation.

#### Conclusions

Preeclampsia in twin pregnancies appears to be associated with biological alterations that are detectable before the onset of clinical disease and may differ from those reported in singleton gestations. These findings provide further evidence that twin pregnancies should not simply be considered an extension of singleton pregnancies when developing risk assessment strategies. Improved understanding of the unique biology underlying twin PE may facilitate the future development of twin-specific approaches for early risk stratification and individualized surveillance.

## GLOSSARY

Preeclampsia: A placenta-mediated hypertensive disorder of pregnancy characterized by new-onset hypertension with maternal organ dysfunction after 20 weeks of gestation. Differentially Expressed Proteins: Proteins exhibiting statistically significant differences in abundance between comparison groups.
KEGG Pathway Analysis: A computational approach using the Kyoto Encyclopedia of Genes and Genomes database to identify enriched biological pathways among protein sets.
Trophoblast Invasion: The process by which extravillous trophoblasts infiltrate the maternal decidua to establish placental perfusion; impaired in preeclampsia.
Ferroptosis: An iron-dependent regulated cell death pathway characterized by lipid peroxidation, implicated in placental oxidative stress.

## Acknowledgments

We sincerely express our gratitude to all the patients who participated in the recruitment process for their generous contribution of their time and samples. We thank the Center for Precision Medicine Multi-Omics Research, Institute of Advanced Clinical Medicine, Peking University for the quantitative mass spectrum analysis.

## Ethical statement

This study was approved by the ethical review board of the Peking University Third Hospital (No. M2023758) and all patients included have signed informed consent.

## Author contributions

This work was a collaborative effort by all authors. J.H., Y.W., and Y.C. conceived and designed the experiments. J.L., Z.F., Q.W., and X.L. conducted the majority of the biochemical experiments, including ELISA, cellular, and histochemical analyses. T. W., Y.F., and J.M. were responsible for data statistics analysis and the establishment of prediction models. W.T. and Y.C. carried out the LC-MS/MS and bioinformatics analyses. J.H., J.L., Z.F., and T.W. wrote and revised the manuscript, with input and feedback from all authors.

## Diversity, equity, ethics, and inclusion

We support inclusive, diverse, and equitable research conduct.

## Data availability statement

The mass spectrometry proteomics data generated in this study have been deposited to the ProteomeXchange Consortium via the PRIDE partner repository with the dataset identifier PXD059849 (http://proteomecentral.proteomexchange.org/cgi/GetDataset?ID=PXD059849), and to the iProX Consortium with the dataset identifier IPX0010798000 (https://www.iprox.cn/page/project.html?id=IPX0010798000).

## References

1. Magee LA, Nicolaides KH, von Dadelszen P. Preeclampsia. N Engl J Med 2022;386:1817–32.

2. Ma’ayeh M, Costantine MM. Prevention of preeclampsia. Semin Fetal Neonatal Med 2020;25:101123.

3. Droge L, Herraiz I, Zeisler H, et al. Maternal serum sFlt-1/PlGF ratio in twin pregnancies with and without pre-eclampsia in comparison with singleton pregnancies. Ultrasound Obstet Gynecol 2015;45:286–93.

4. Yang M, Bai Y, Li M, Lin X, Duan X, Zhang X. Predictive value of the soluble fms-like tyrosine kinase 1 to placental growth factor ratio for preeclampsia in twin pregnancies: a systematic review and meta-analysis. Am J Obstet Gynecol MFM 2024;6:101290.

5. Stepan H, Galindo A, Hund M, et al. Clinical utility of sFlt-1 and PlGF in screening, prediction, diagnosis and monitoring of pre-eclampsia and fetal growth restriction. Ultrasound Obstet Gynecol 2023;61:168–80.

6. Saleh L, Tahitu SIM, Danser AHJ, van den Meiracker AH, Visser W. The predictive value of the sFlt-1/PlGF ratio on short-term absence of preeclampsia and maternal and fetal or neonatal complications in twin pregnancies. Pregnancy Hypertens 2018;14:222–27.

7. Magee LA, Brown MA, Hall DR, et al. The 2021 International Society for the Study of Hypertension in Pregnancy classification, diagnosis & management recommendations for international practice. Pregnancy Hypertens 2022;27:148–69.

8. Bdolah Y, Lam C, Rajakumar A, et al. Twin pregnancy and the risk of preeclampsia: bigger placenta or relative ischemia? Am J Obstet Gynecol 2008;198:428.e1–6.

9. Rana S, Hacker MR, Modest AM, et al. Circulating angiogenic factors and risk of adverse maternal and perinatal outcomes in twin pregnancies with suspected preeclampsia. Hypertension 2012;60:451–8.

10. Avnon T, Ascher-Landsberg J, Yogev Y, Many A. Clinical characteristics of preeclampsia in twin versus singleton pregnancies. J Matern Fetal Neonatal Med 2022;35:2273–77.

11. Aviram A, Berger H, Abdulaziz KE, et al. Outcomes Associated With Hypertensive Disorders of Pregnancy in Twin Compared With Singleton Gestations. Obstet Gynecol 2021;138:449–58.

12. Giorgione V, Melchiorre K, O’Driscoll J, Khalil A, Sharma R, Thilaganathan B. Maternal echocardiographic changes in twin pregnancies with and without pre-eclampsia. Ultrasound Obstet Gynecol 2022;59:619–26.

13. Elkayam U, Akhter MW, Singh H, et al. Pregnancy-associated cardiomyopathy: clinical characteristics and a comparison between early and late presentation. Circulation 2005;111:2050–5.

14. Thilaganathan B. Pre-eclampsia and the cardiovascular-placental axis. Ultrasound Obstet Gynecol 2018;51:714–17.

15. Jing W, Gu X, Yang J, Wei Y, Zhao Y. Maternal lipid levels in preeclampsia: singleton vs. twin pregnancies. J Matern Fetal Neonatal Med 2022;35:6132–39.

16. Erez O, Vardi IS, Hallak M, Hershkovitz R, Dukler D, Mazor M. Preeclampsia in twin gestations: association with IVF treatments, parity and maternal age. J Matern Fetal Neonatal Med 2006;19:141–6.

17. Hiersch L, Berger H, Okby R, et al. Gestational diabetes mellitus is associated with adverse outcomes in twin pregnancies. Am J Obstet Gynecol 2019;220:102.e1–02.e8.

18. Aghaeepour N, Lehallier B, Baca Q, et al. A proteomic clock of human pregnancy. Am J Obstet Gynecol 2018;218:347 e1–47 e14.

19. Romero R, Erez O, Maymon E, et al. The maternal plasma proteome changes as a function of gestational age in normal pregnancy: a longitudinal study. Am J Obstet Gynecol 2017;217:67 e1–67 e21.

20. Ho SJ, Chaput D, Sinkey RG, et al. Proteomic studies of VEGFR2 in human placentas reveal protein associations with preeclampsia, diabetes, gravidity, and labor. Cell Commun Signal 2024;22:221.

21. Lim JH, Lim JM, Lee HM, et al. Systematic Proteome Profiling of Maternal Plasma for Development of Preeclampsia Biomarkers. Mol Cell Proteomics 2024;23:100826.

22. Lindley KJ, Perry A, Jacobs M, et al. Differences in Cardiometabolic Proteins in Pregnancy Prioritize Relevant Targets of Preeclampsia. Arterioscler Thromb Vasc Biol 2024;44:969–75.

23. Tarca AL, Romero R, Benshalom-Tirosh N, et al. The prediction of early preeclampsia: Results from a longitudinal proteomics study. PLoS One 2019;14:e0217273.

24. Tarca AL, Taran A, Romero R, et al. Prediction of preeclampsia throughout gestation with maternal characteristics and biophysical and biochemical markers: a longitudinal study. Am J Obstet Gynecol 2022;226:126 e1–26 e22.

25. Burwick RM, Feinberg BB. Complement activation and regulation in preeclampsia and hemolysis, elevated liver enzymes, and low platelet count syndrome. Am J Obstet Gynecol 2022;226:S1059–s70.

26. Stincone A, Prigione A, Cramer T, et al. The return of metabolism: biochemistry and physiology of the pentose phosphate pathway. Biol Rev Camb Philos Soc 2015;90:927–63.

27. Wang S, Liu X, Meng Z, et al. DCBLD2 regulates vascular hyperplasia by modulating the platelet derived growth factor receptor-β endocytosis through Caveolin-1 in vascular smooth muscle cells. Faseb j 2022;36:e22488.

28. Tan MY, Syngelaki A, Poon LC, et al. Screening for pre-eclampsia by maternal factors and biomarkers at 11-13 weeks’ gestation. Ultrasound Obstet Gynecol 2018;52:186–95.

29. MacDonald TM, Walker SP, Hannan NJ, Tong S, Kaitu’u-Lino TJ. Clinical tools and biomarkers to predict preeclampsia. EBioMedicine 2022;75:103780.

30. Hamdan HZ. Exploring gene expression signatures in preeclampsia and identifying hub genes through bioinformatic analysis. Placenta 2025;159:93–106.

31. Lim HI, Hajjar KA. Annexin A2 in Fibrinolysis, Inflammation and Fibrosis. Int J Mol Sci 2021;22.

32. Maxwell CV, Lieberman E, Norton M, Cohen A, Seely EW, Lee-Parritz A. Relationship of twin zygosity and risk of preeclampsia. Am J Obstet Gynecol 2001;185:819–21.

33. Savvidou MD, Karanastasi E, Skentou C, Geerts L, Nicolaides KH. Twin chorionicity and pre-eclampsia. Ultrasound Obstet Gynecol 2001;18:228–31.

34. Dong J, Hu Y, Liu S, et al. Arsenic induces ferroptosis in HTR-8/SVneo cells and placental damage. Sci Total Environ 2024;955:176885.

35. Perucci LO, Carneiro FS, Ferreira CN, et al. Annexin A1 Is Increased in the Plasma of Preeclamptic Women. PLoS One 2015;10:e0138475.

36. Feng J, Wang X, Li H, Wang L, Tang Z. Silencing of Annexin A1 suppressed the apoptosis and inflammatory response of preeclampsia rat trophoblasts. Int J Mol Med 2018;42:3125–34.

37. Schrader JM, Stanisavljevic A, Xu F, Van Nostrand WE. Distinct Brain Proteomic Signatures in Cerebral Small Vessel Disease Rat Models of Hypertension and Cerebral Amyloid Angiopathy. J Neuropathol Exp Neurol 2022;81:731–45.

38. Cañas F, Simonin L, Couturaud F, Renaudineau Y. Annexin A2 autoantibodies in thrombosis and autoimmune diseases. Thromb Res 2015;135:226–30.

39. Hu J, Chen L, Ruan J, Chen X. The role of the annexin A protein family at the maternal-fetal interface. Front Endocrinol (Lausanne) 2024;15:1314214.

40. Sano M, Matsumoto M, Terada H, et al. Increased annexin A2 expression in the placenta of women with acute worsening of preeclampsia. Osaka City Med J 2014;60:87–93.

41. Garrido-Gomez T, Quiñonero A, Dominguez F, et al. Preeclampsia: a defect in decidualization is associated with deficiency of Annexin A2. Am J Obstet Gynecol 2020;222:376.e1–76.e17.

42. Xin H, Zhang Y, Wang H, Sun S. Alterations of profibrinolytic receptor annexin A2 in pre- eclampsia: a possible role in placental thrombin formation. Thromb Res 2012;129:563–7.

43. Abd El-Latif M, Azzam H, Othman M, Warda O, El-Sharawy S, Ghoneim H. Assessment of annexin A5 and annexin A2 levels as biomarkers for pre-eclampsia: A pilot study. Pregnancy Hypertens 2017;8:65–69.

44. Zhang N, Yu X, Xie J, Xu H. New Insights into the Role of Ferritin in Iron Homeostasis and Neurodegenerative Diseases. Mol Neurobiol 2021;58:2812–23.

45. Yang X, Ding Y, Sun L, et al. Ferritin light chain deficiency-induced ferroptosis is involved in preeclampsia pathophysiology by disturbing uterine spiral artery remodelling. Redox Biol 2022;58:102555.

