## Supplemental Data for "Early maternal plasma protein signatures identify preeclampsia risk in twin pregnancies"

Supplemental Figures 1 to 10

Supplemental Tables 1 to 3

Supplemental materials and methods

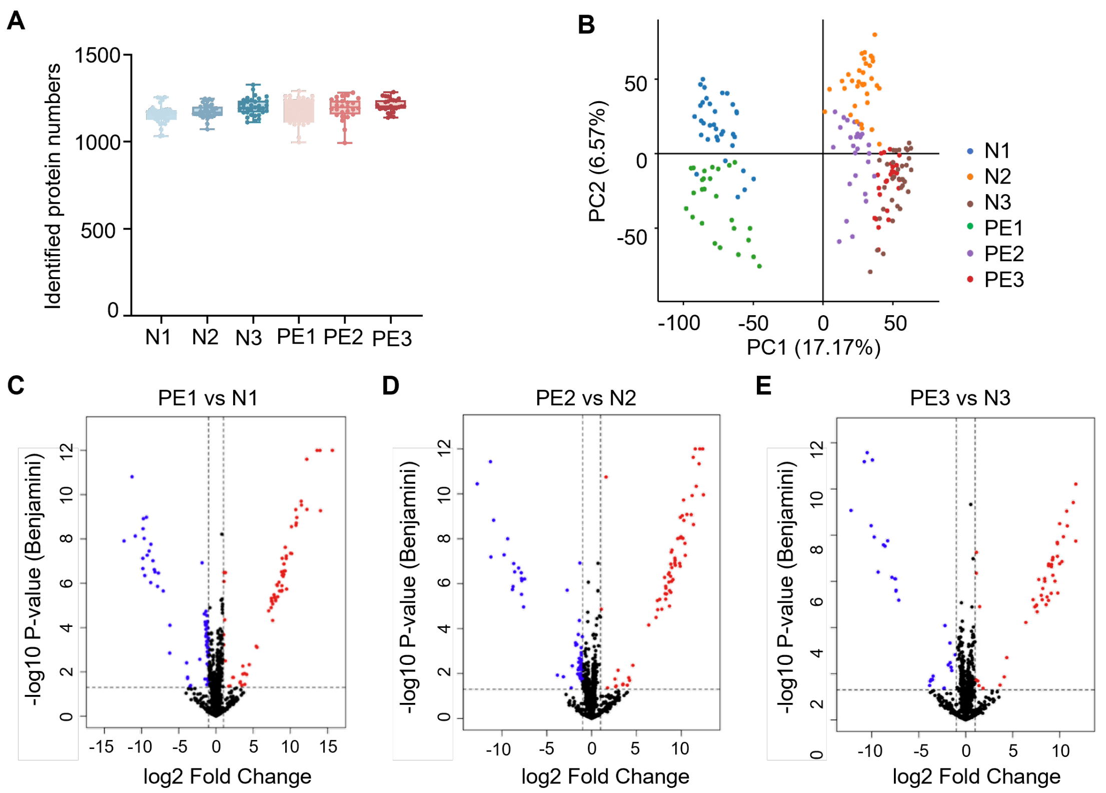


**Supplemental Figure 1. Quality control of the proteomic profiling.** (**A**) Numbers of proteins identified at each gestational stage. N1, N2, and N3: first, second, and third trimesters in the healthy normotensive group; PE1, PE2, and PE3: first, second, and third trimesters in the twin PE group. (**B**) Principal component analysis (PCA) of samples with all detected proteins across the six gestational stages. (**C–E**) Volcano plot of differentially expressed proteins (DEPs) between PE and N in the first (PE1 vs N1, **C**), second (PE2 vs N2, **D**), and third (PE3 vs N3, **E**) trimesters. The p-values were adjusted using the Benjamini-Hochberg multiple testing correction. Statistically significant DEPs were represented in blue (downregulated) and red (upregulated).

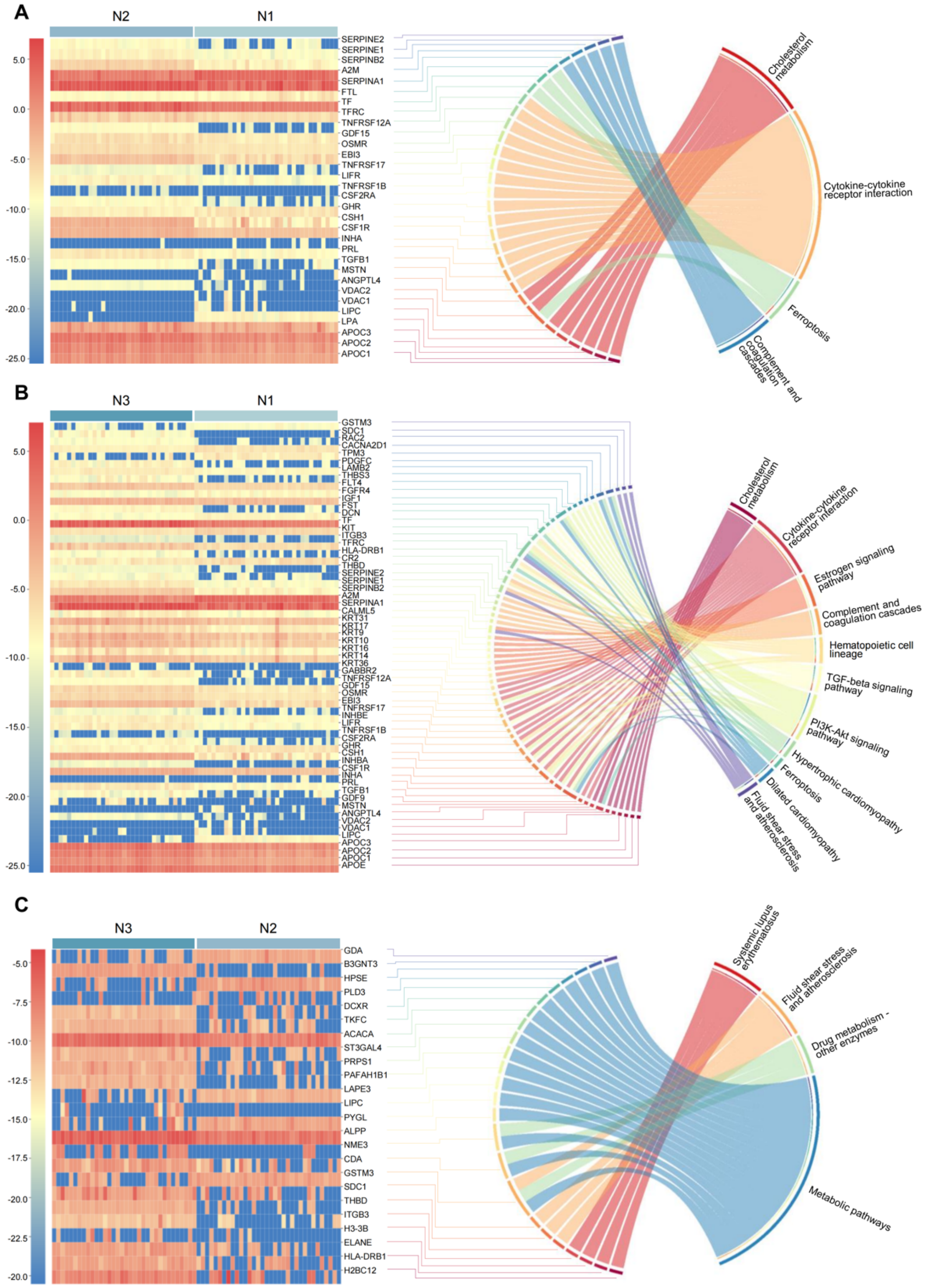


**Supplemental Figure 2. Trimester-specific differentially expressed proteins (DEPs) and KEGG pathway enrichment in healthy control twin pregnancy.** (**A**–**C**) Heatmaps and chord diagrams demonstrating DEPs and the corresponding enriched pathways between N2 and N1 (**A**), N3 and N1 (**B**), and N3 and N2 (**C**).

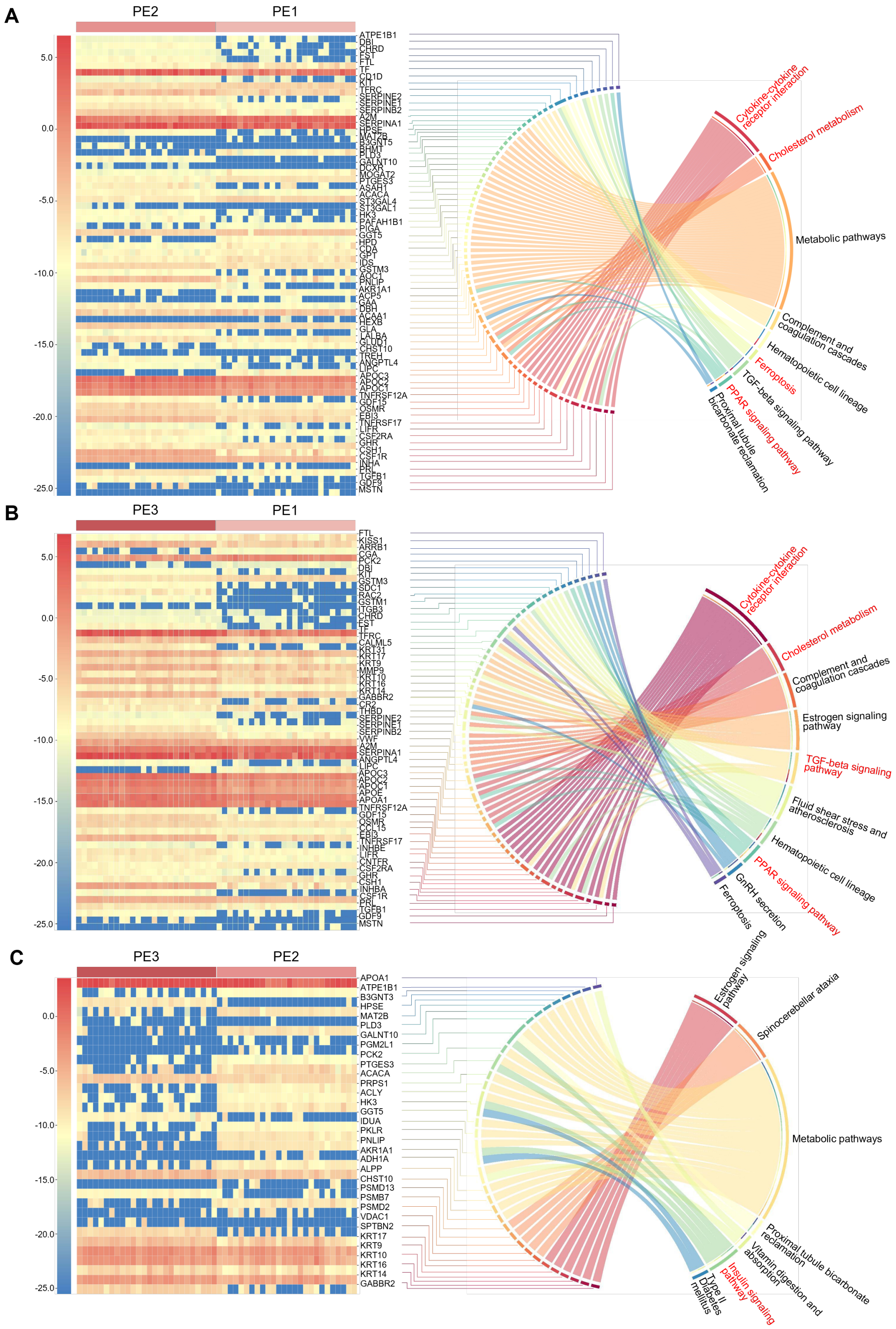


**Supplemental Figure 3. Gestational Stage-Specific Proteomic Alterations in Twin PE across three trimesters.** (**A–C**) Heatmaps and chord diagrams demonstrating the DEPs and enriched pathways between PE2 and PE1 (A), PE3 and PE1 (B), and PE3 and PE2 (C).

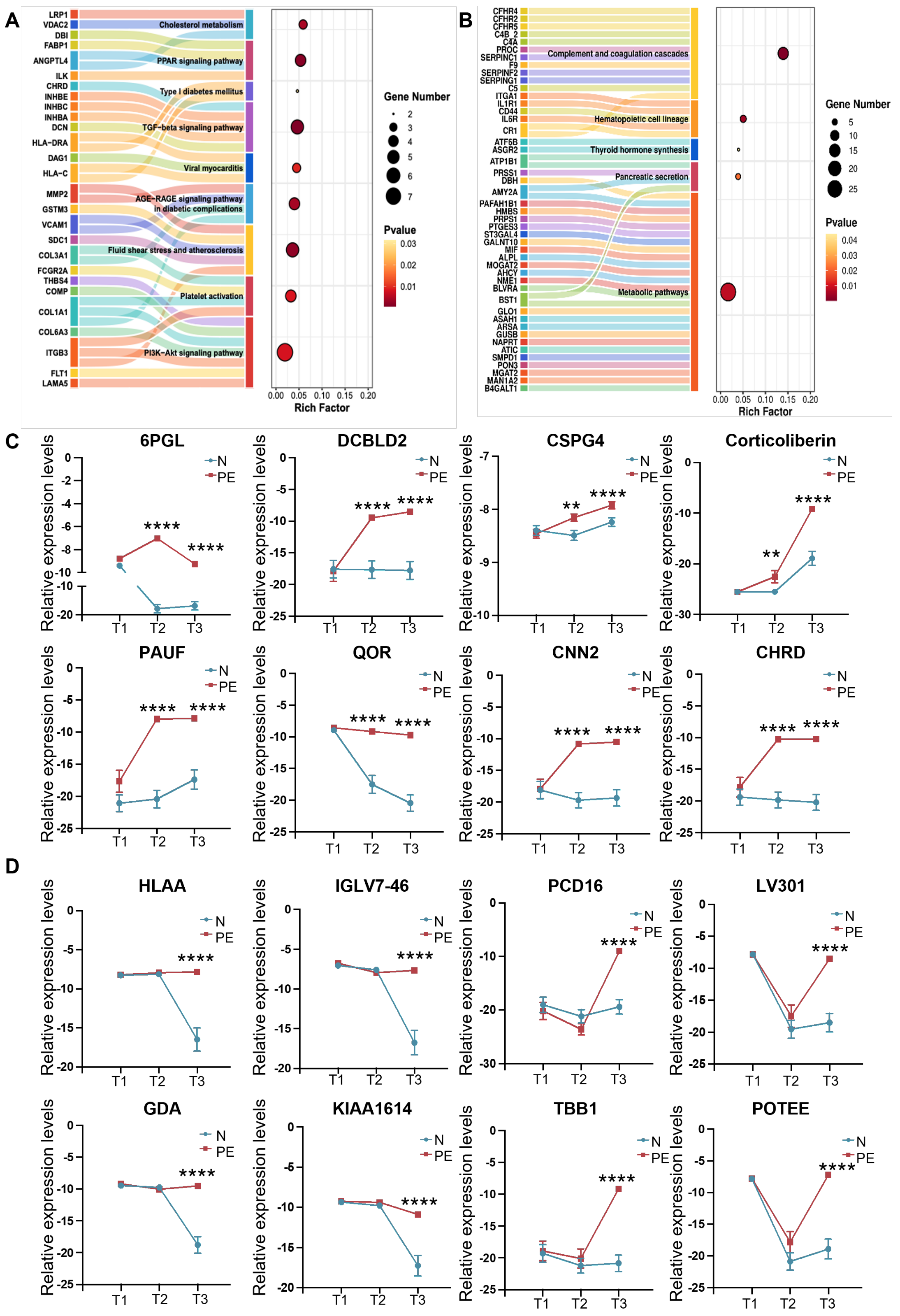


**Supplemental Figure 4. Proteins exhibiting divergent expression trends between PE and N groups.** (**A–B**) KEGG pathway enrichment analyses of proteins upregulated (**A**) or downregulated (**B**) in the second and third trimester. (**C**) Representative expression profiles of selected proteins increasing in the second and third trimester. 6PGL, 6-phosphogluconolactonase; DCBLD2, discoidin, CUB and LCCL domain-containing protein 2; CSPG4, chondroitin sulfate proteoglycan 4; Corticoliberin; PAUF, pancreatic adenocarcinoma up-regulated factor; QOR, quinone oxidoreductase; CNN2, calponin-2; CHRD, chordin; (**D**) Representative expression profiles of selected proteins increasing in the third trimester. HLA-A, HLA class I histocompatibility antigen, A alpha chain; IGLV7-46, immunoglobulin lambda variable 7-46; PCD16, protocadherin-16; LV301, immunoglobulin lambda variable 3-1; GDA, guanine deaminase; KIAA1614, uncharacterized protein KIAA1614; TBB1, tubulin beta-1 chain; POTEE, POTE ankyrin domain family member E. Two-way ANOVA was used for statistical analysis. ** $P < 0.01$, **** $P < 0.0001$.

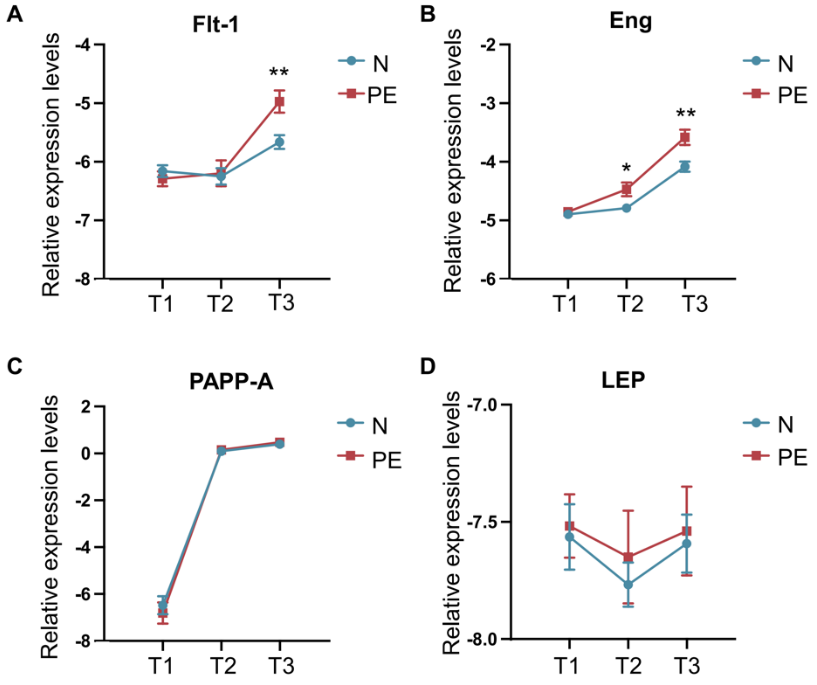


**Supplemental Figure 5. Proteomic expression profiles of classical predictive biomarkers.** (**A–B**) Maternal plasma expression profiles of fms-like tyrosine kinase-1 (Flt-1, **A**), Endoglin (Eng, **B**), pregnancy-associated plasma protein A (PAPP-A, **C**), and leptin (LEP, **D**) across T1, T2, and T3 in PE and N groups. Two-way ANOVA was used for statistical analysis. * $P < 0.05$, ** $P < 0.01$.

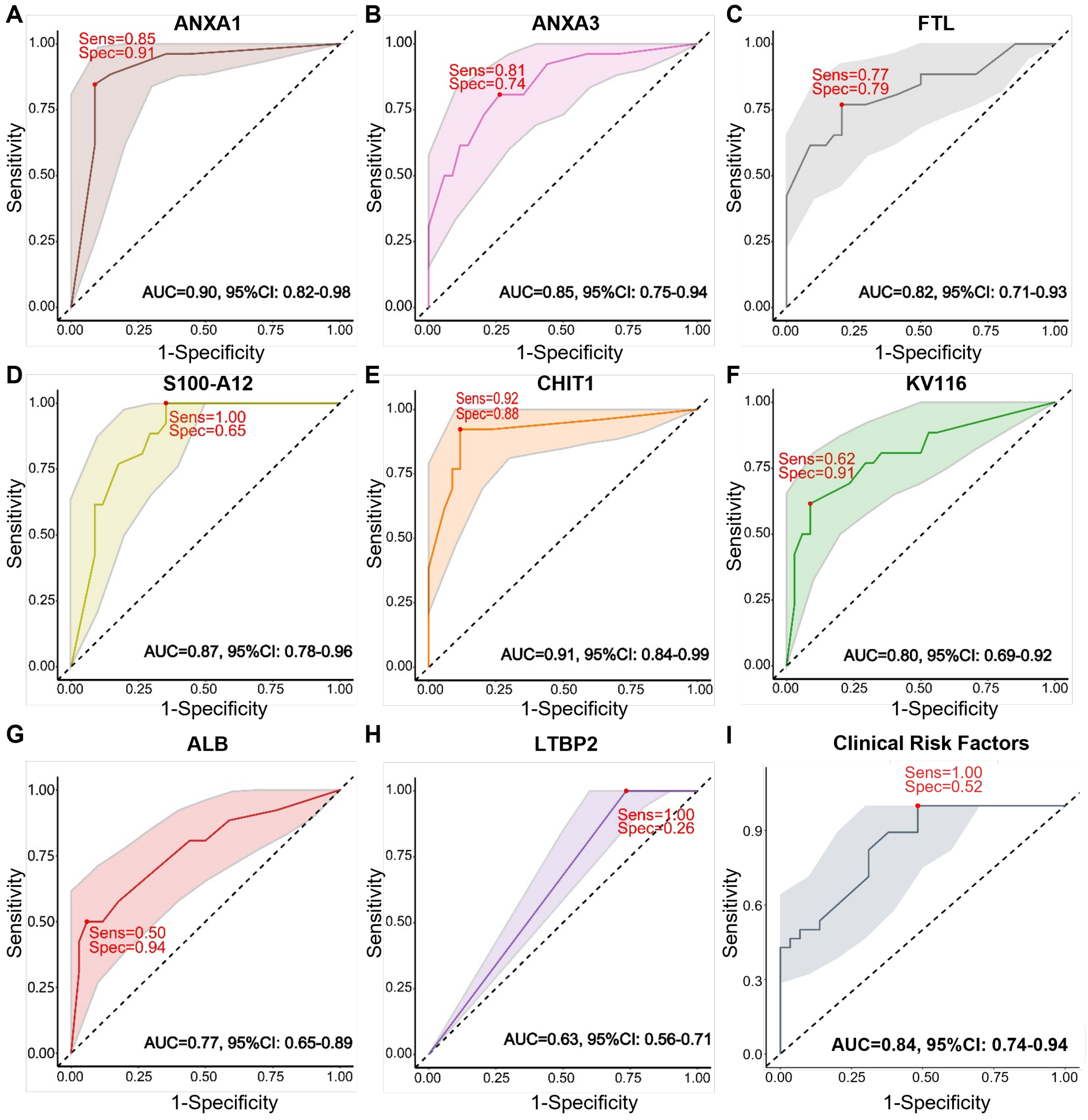


**Supplemental Figure 6. XGBoost machine learning-based early predictive models.** (**A–I**) An XGBoost machine learning-based early predictive model utilizes ANXA1 (**A**), ANXA3 (**B**), FTL (**C**), S100-A12 (**D**), CHIT1 (**E**), KV116 (**F**), ALB (**G**), LTBP2 (**H**) and clinical risk factors (**I**). Sens, sensitivity. Spec, specificity.

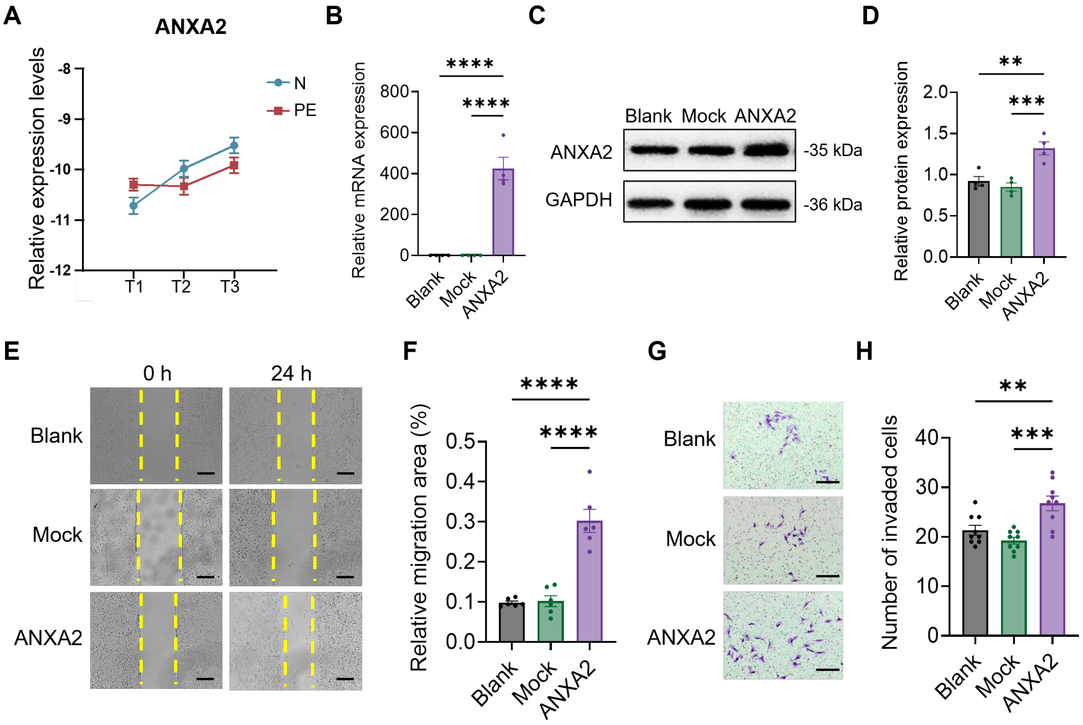


**Supplemental Figure 7. Plasma annexin A2 expression and functional characterization of ANXA2 overexpression in trophoblasts.** (**A**) Protein expression profile of Annexin A2 (ANXA2) across three trimesters determined by proteomics. T1, T2, and T3, first-, second-, and third-trimesters. (**B–D**) ANXA2 overexpression in HTR8/SVneo trophoblast cells conformed by RT-qPCR (**B**) and Western blot (**C, D**). Data represents four independent biological replicates. Blank, no transfected; mock, empty vector; ANXA2, ANXA2 overexpression. (**E–F**) Migration assay of HTR8/SVneo cells after ANXA2 overexpression (**E**) and the corresponding quantitative assessment based on scratch closure area (**F**). Scale bars: 250 μm. (**G–H**) Invasion assay for ANXA2 overexpression (**G**) and the corresponding quantitative assessment based on the number of invading cells (**H**). Scale bars: 50 μm. ** $P < 0.01$, *** $P < 0.001$, **** $P < 0.0001$.

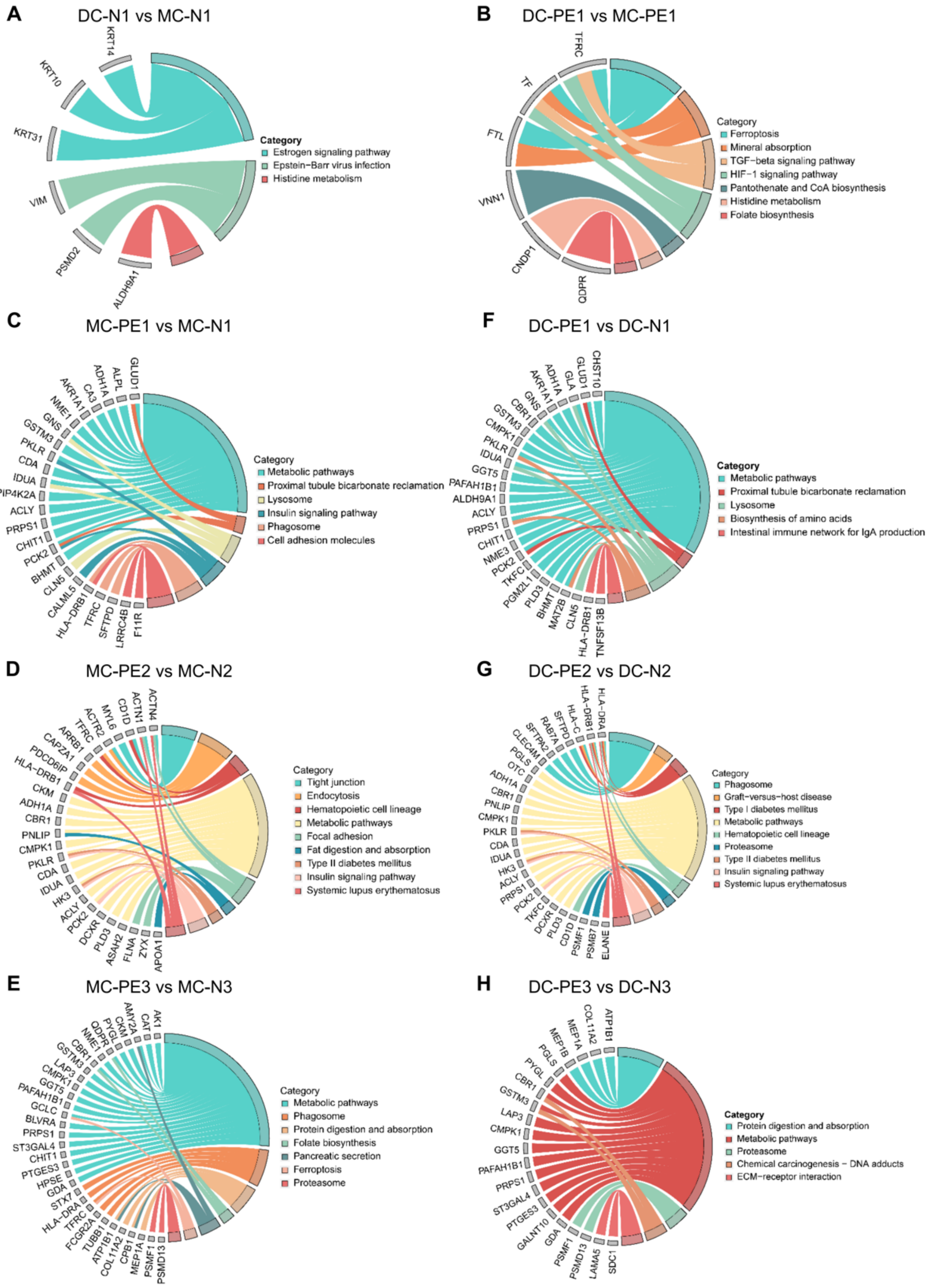


**Supplemental Figure 8. Monochorionic (MC) or dichorionic (DC)-specific DEPs and KEGG pathway enrichment in twin PE and normotensive groups.** (**A–H**) Chord diagrams demonstrating the DEPs and enriched pathway between DC-N1 vs MC-N1 (**A**), DC-PE1 vs MC-PE1 (**B**), MC-PE1 vs MC-N1 (**C**), MC-PE2 vs MC-N2 (**D**), MC-PE3 vs MC-N3 (**E**), DC-PE1 vs DC-N1 (**F**), DC-PE2 vs DC-N2 (**G**), and DC-PE3 vs DC-N3 (**H**).

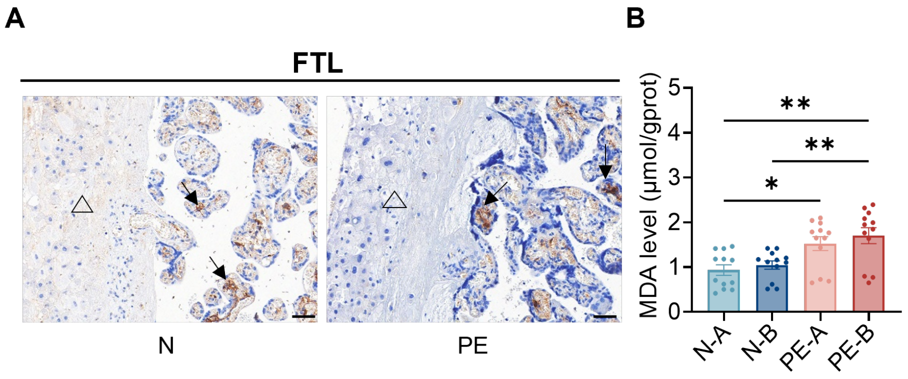


**Supplemental Figure 9. Oxidative status of placental tissues in PE versus N groups.** (**A**) Immunohistochemistry (IHC) staining of FTL expression in placental villi (indicated by arrows) and decidua (indicated by triangles). Scale bars = 50 μm. (**B**) Malondialdehyde (MDA) levels in placental villi. N-A and N-B, twin placental shares derived from normotensive twin pregnancies. PE-A and PE-B, twin placental shares from the twin PE pregnancies. Four independent placental shares were used for both PE and N groups with one-way ANOVA statistics. * $P < 0.05$, ** $P < 0.01$.

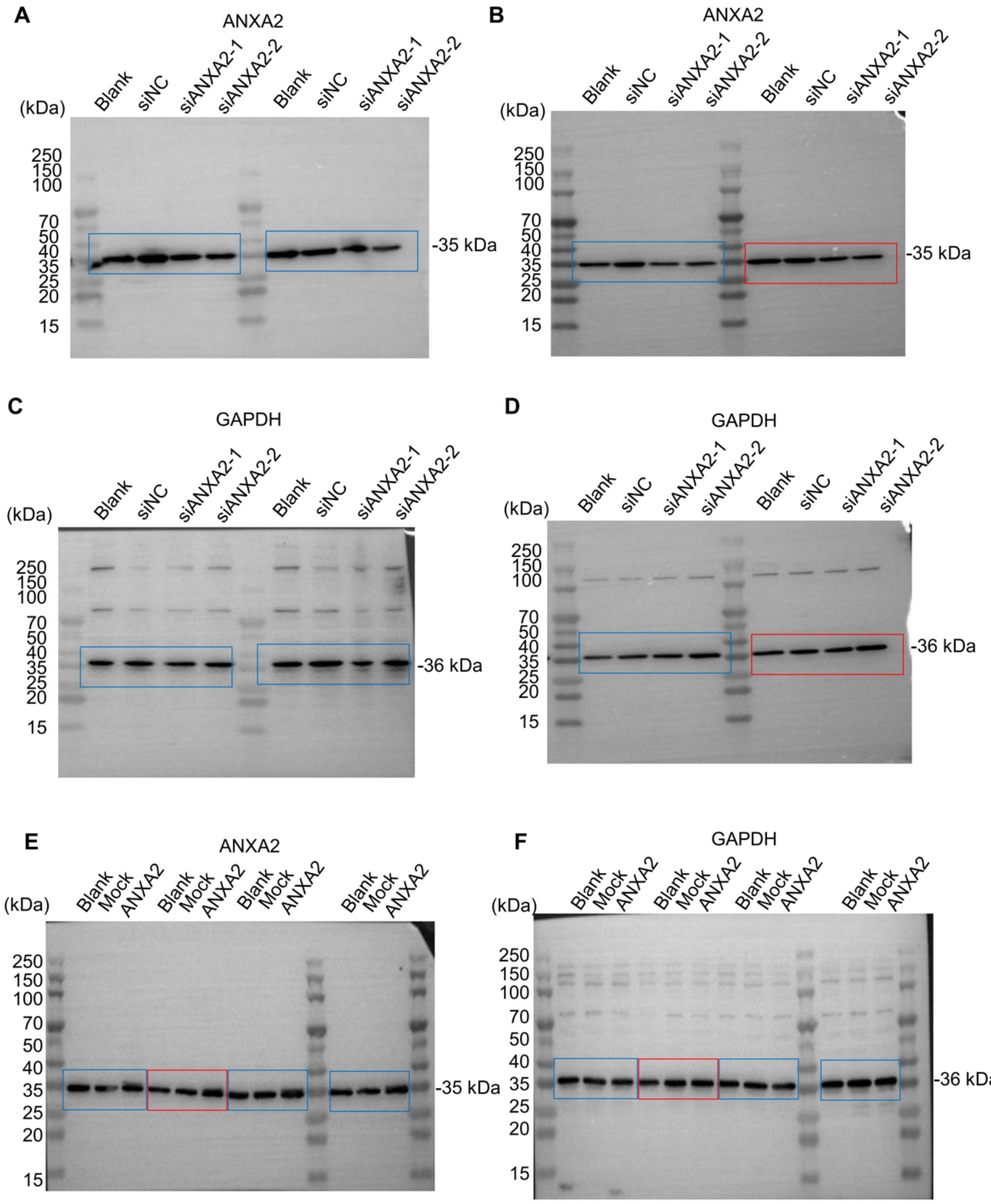


**Supplemental Figure 10. Uncropped gels for Western blots.** (**A–D**) Knockdown of ANXA2 in the HTR8/SVneo cells. ANXA2 (**A, B**) and GAPDH as loading control (**C, D**) were separately blotted. The red boxes indicate the presented bands in Figure 7C and the blue boxes used for statistics. (**E–F**) ANXA2 overexpression in the HTR8/SVneo cells. ANXA2 (**E**) and GAPDH as loading control (**F**) with red boxes in Figure S7B and blue boxes for statistics.

**Supplemental Table 1. Performance of different machine learning algorithms for early prediction of twin PE**

| Early differential proteins | Model | AUC | CI_lower | CI_upper |
|---|---|---|---|---|
| ANXA1 | Logistic | 0.844 | 0.73 | 0.957 |
| | DecisionTree | 0.869 | 0.782 | 0.956 |
| | NaiveBayes | 0.922 | 0.84 | 1 |
| | SVM | 0.932 | 0.868 | 0.997 |
| | XGBoost | 0.901 | 0.818 | 0.984 |
| | KNN | 0.932 | 0.873 | 0.99 |
| ANXA3 | Logistic | 0.739 | 0.609 | 0.868 |
| | DecisionTree | 0.771 | 0.664 | 0.879 |
| | NaiveBayes | 0.739 | 0.609 | 0.868 |
| | SVM | 0.817 | 0.707 | 0.926 |
| | XGBoost | 0.846 | 0.746 | 0.945 |
| | KNN | 0.811 | 0.702 | 0.919 |
| FTL | Logistic | 0.665 | 0.519 | 0.811 |
| | DecisionTree | 0.68 | 0.558 | 0.801 |
| | NaiveBayes | 0.721 | 0.592 | 0.849 |
| | SVM | 0.741 | 0.614 | 0.868 |
| | XGBoost | 0.821 | 0.709 | 0.934 |
| | KNN | 0.708 | 0.578 | 0.838 |
| S100-A12 | Logistic | 0.679 | 0.537 | 0.821 |
| | DecisionTree | 0.733 | 0.618 | 0.848 |
| | NaiveBayes | 0.84 | 0.741 | 0.94 |
| | SVM | 0.904 | 0.827 | 0.981 |
| | XGBoost | 0.872 | 0.783 | 0.961 |
| | KNN | 0.843 | 0.746 | 0.939 |
| CHIT1 | Logistic | 0.874 | 0.779 | 0.97 |
| | DecisionTree | 0.859 | 0.772 | 0.945 |
| | NaiveBayes | 0.871 | 0.773 | 0.969 |
| | SVM | 0.925 | 0.84 | 1 |
| | XGBoost | 0.914 | 0.835 | 0.993 |
| | KNN | 0.932 | 0.876 | 0.987 |
| KV116 | Logistic | 0.692 | 0.558 | 0.826 |
| | DecisionTree | 0.764 | 0.649 | 0.879 |
| | NaiveBayes | 0.692 | 0.558 | 0.826 |
| | SVM | 0.812 | 0.706 | 0.918 |
| | XGBoost | 0.802 | 0.685 | 0.919 |
| | KNN | 0.764 | 0.649 | 0.879 |
| ALB | Logistic | 0.683 | 0.544 | 0.822 |
| | DecisionTree | 0.801 | 0.687 | 0.915 |
| | NaiveBayes | 0.658 | 0.513 | 0.804 |

| | | | | |
|---|---|---|---|---|
| | SVM | 0.218 | 0.1 | 0.337 |
| | XGBoost | 0.77 | 0.647 | 0.893 |
| | KNN | 0.773 | 0.657 | 0.889 |
| LTBP2 | Logistic | 0.534 | 0.414 | 0.654 |
| | DecisionTree | 0.731 | 0.64 | 0.822 |
| | NaiveBayes | 0.534 | 0.414 | 0.654 |
| | SVM | 0.731 | 0.64 | 0.822 |
| | XGBoost | 0.632 | 0.557 | 0.708 |
| | KNN | 0.731 | 0.64 | 0.822 |
| CHIT1+ANXA1+ANXA3+FTL+S100-A12 | Logistic | 0.957 | 0.903 | 1 |
| | DecisionTree | 0.928 | 0.856 | 1 |
| | NaiveBayes | 0.95 | 0.895 | 1 |
| | SVM | 0.971 | 0.921 | 1 |
| | XGBoost | 0.963 | 0.918 | 1 |
| | KNN | 0.971 | 0.934 | 1 |

**Supplemental Table 2. Clinical information of placental shares used for immunohistochemistry (IHC).**

| | N (n=4) | PE (n=4) | *P* value |
|---|---|---|---|
| Maternal age, median (IQR), y | 33.50 (30.75~37.00) | 34.00 (29.50~37.00) | 0.882 |
| Pre-pregnancy weight, median (IQR), kg | 56.50 (51.88~66.00) | 61.50 (50.63~64.50) | 0.885 |
| Pre-pregnancy BMI, median (IQR), kg/m$^2$ | 21.60 (21.08~23.30) | 21.79 (18.85~24.26) | 0.686 |
| Parity, n (%) | | | 0.285 |
| Primipara | 3 (75.00) | 4 (100.00) | |
| Multipara | 1 (25.00) | 0 (00.00) | |
| Conception method, n (%) | | | 0.465 |
| Spontaneous conception | 3 (75.00) | 2 (50.00) | |
| Assisted reproductive techniques | 1 (25.00) | 2 (50.00) | |
| Chorionicity, n (%) | | | 0.757 |
| Monochorionic | 2 (50.00) | 2 (50.00) | |
| Dichorionic | 2 (50.00) | 2 (50.00) | |
| Nation, n (%) | | | |
| Han Chinese | 3 (75.00) | 3 (75.00) | 0.786 |
| Ethnic Minorities | 1 (25.00) | 1 (25.00) | |
| Mean Arterial Pressure at 11~14 weeks of gestation, median (IQR), mmHg | 97.67 (80.17~101.17) | 88.83 (86.33~103.33) | 0.886 |
| Chronic hypertension complicating pregnancy, n (%) | 1 (25.00) | 1 (25.00) | 0.786 |
| Pregestational diabetes mellitus, n (%) | 0 (00.00) | 0 (00.00) | |
| Autoimmune disease complicating pregnancy, n (%) | 0 (00.00) | 0 (00.00) | |
| Gestational age at delivery, median (IQR), week | $37^{+0}$ ($34^{+0}$~$37^{+2}$) | $36^{+0}$ ($35^{+1}$~$37^{+0}$) | 0.029 |
| Gestational diabetes mellitus, n (%) | 0 (00.00) | 2 (50.00) | 0.102 |
| Birth weight, median (IQR), kg | | | |
| The fetus above | 2500.00 (2355.00~2885.00) | 2030.00 (1402.50~2455.00) | 0.114 |
| Birth weight < $10^{th}$ | 2 (50.00%) | 2 (50.00%) | 0.757 |
| Birth weight > $90^{th}$ | 0 (0.00%) | 0 (0.00%) | / |
| The fetus below | 2485.00 (2225.00~2760.00) | 2165.00 (1440.00~2942.50) | 0.686 |
| Birth weight < $10^{th}$ | 2 (50.00%) | 2 (50.00%) | 0.757 |

| | | | |
|---|---|---|---|
| Birth weight > 90$^{th}$ | 0 (0.00%) | 0 (0.00%) | / |
| Neonatal intensive care units admission, n (%) | 1 (25.00) | 1 (25.00) | 0.786 |

IQR, Interquartile range; BMI, Body mass index; The fetus above, the twin delivered first; The fetus below, the twin delivered second.

**Supplemental Table 3. Comparison of prediction model performance between clinical risk factors and combined biomarkers using different machine learning algorithms for twin PE.**

| Early differential proteins | Model | AUC | CI_lower | CI_upper |
|---|---|---|---|---|
| Clinical risk factors | Logistic | 0.688 | 0.548 | 0.829 |
| | DecisionTree | 0.700 | 0.581 | 0.819 |
| | NaiveBayes | 0.826 | 0.713 | 0.940 |
| | SVM | 0.738 | 0.607 | 0.868 |
| | XGBoost | 0.842 | 0.743 | 0.940 |
| | KNN | 0.740 | 0.614 | 0.866 |
| ANXA2+ANXA3+CHIT1+FTL+Clinical risk factors | Logistic | 0.875 | 0.744 | 1 |
| | DecisionTree | 0.740 | 0.569 | 0.910 |
| | NaiveBayes | 0.948 | 0.869 | 1 |
| | SVM | 0.984 | 0.953 | 1.000 |
| | XGBoost | 0.974 | 0.929 | 1.000 |
| | KNN | 0.807 | 0.639 | 0.976 |

### Supplemental Materials and Methods

***Participants and Sample Collection***: This nested case-control study was conducted within the prospective twin birth cohort established by the Department of Obstetrics at Peking University Third Hospital. The study was approved by thePeking University Third Hospital Medical Science Research Ethics Committee (No. M2023758). The parent cohort prospectively recruited women aged 18–45 with twin pregnancies (< 14 weeks gestation) between 2023 and 2024. All participants provided written informed consent, underwent standardized prenatal examinations, and delivered at the research center. From this cohort, we identified 40 incident cases of PE in twin pregnancies, which was defined according to International Society for the Study of Hypertension in Pregnancy (ISSHP)[53] criteria. Cases were confirmed by independent medical record review. Controls were matched to cases based on maternal age, pre-pregnancy weight, pre-pregnancy BMI, parity, conception method, chorionicity. Pregnancies complicated by selective fetal growth restriction (sFGR), twin-twin transfusion syndrome (TTTS), twin reversed arterial perfusion sequence (TRAPS), or twin anemia-polycythemia sequence (TAPS) were excluded. Maternal blood samples were collected at predetermined time points in the first (N1, PE1), second (N2, PE2), and third (N3, PE3) trimesters alongside routine prenatal assessments. Plasma was processed and stored at -80°C until analysis. For this study, biospecimens from the selected case-control pairs were retrieved for batch analysis, with laboratory personnel blinded to case-control status.

***Depletion of High-Abundance Proteins***: Maternal plasma (20 μL) was thoroughly mixed with 60 μL Buffer A (Agilent, 5185-5987), transferred into a 0.22 μm cellulose acetate filter-certified tube (Costor), and centrifuged at 10,000 × *g* for 1 min. The filtrate was applied to 29 × 5 mm High-Select Top14 Abundant Protein Depletion column (Agilent, 51886557) operated on Agilent Technologies 1290 LC system. The depleted samples were collected, and the protein concentration was quantified using a BCA Protein Assay Kit (Thermo Scientific, 23225) following the manufacturer's protocol.

***Sample Preparation for LC-MS/MS***: The high-abundance protein-depleted plasma sample (50 μg) was precipitated with 1/3 volume of trichloroacetic acid (TCA) solution at 4 °C overnight, followed by centrifugation at 16,000 × *g* for 30 min at 4 °C. The resulting precipitates were washed three times with ice-cold acetone and dried using a vacuum concentrator (Labconco, USA). The dried pellet was dissolved in 40 μL of 8 M urea (pH 8.5), reduced with 20 mM (2-carboxyethyl) phosphine hydrochloride (TCEP), and alkylated with 40 mM iodoacetamide (IAA). After dilution to a final urea concentration of 1.3 M using 200 μL of 100 mM Tris-HCl (pH 8.5), trypsin digestion was carried out with 3 μg of trypsin protease (Promega, V5113) at 37 °C for 16 hours. The digested peptides were desalted using a Monospin C18 column (GL Science, Tokyo, Japan), dried using a vacuum concentrator, and reconstituted in Milli-Q water containing 0.1% formic acid (FA). Indexed retention time (iRT) calibration peptides were added to the samples prior to data-independent acquisition (DIA) analysis.

***LC-MS/MS Analysis for Low-Abundance Protein:*** The LC-MS/MS analysis was carried out using an EASY-nLC 1200 HPLC system (Thermo Scientific, USA) coupled to a Q Exactive HF-X mass spectrometer (Thermo Scientific, USA). Peptide separation was performed on a reversed-phase C18 column (25 cm × 75 μm, 1.7 μm, IonOpticks) over 90 min. Mobile phase A contained 0.1% formic acid in water, and the elution gradient of mobile phase B (0.1% formic acid in 80% acetonitrile) was linearly increased from 9% to 27% over 70 min, then to 45% over 8 min, and to 95% over 5 min, followed by an additional 7 min of equilibration. The flow rate was set to 300 nL/min, and the column temperature was maintained at 50°C. For single-shot proteomics using DIA, the MS1 scan resolution was set to 60,000, with a scan range of 350−1200 m/z. The automatic gain control (AGC) target was set to $3 \times 10^6$, and the maximum injection time was 50 ms. Precursors were isolated with a dynamic isolation width spanning 350−1200 m/z and fragmented using higher-energy collisional dissociation (HCD) at a normalized collision energy (NCE) of 28. MS2 scans were acquired in the orbitrap at a resolution of 30,000, with the maximum injection time set to auto and the AGC target set to $1 \times 10^6$.

***Database Search and Quantification**:* The direct DIA analysis was conducted using Spectronaut version 17.6 (Biognosys) in default mode. Briefly, carbamidomethylation was set as a fixed modification, and acetylation of protein N-termini and oxidation of methionines were set as variable modifications. Trypsin/P was specified as the proteolytic enzyme, allowing a maximum of two missed cleavages. Peptides were filtered to have a minimum length of 7 amino acids and a maximum length of 52 amino acids. The false discovery rate (FDR) was controlled at 1% for both peptide spectra match (PSM) identification and protein quantification.

***Bioinformatics Analysis**:* Differentially expressed proteins (DEPs) were identified using |fold change (FC)| $\geqslant$ 2.0 and $p$-value < 0.05. Principal component analysis (PCA) and Pearson's correlation coefficient were applied to evaluate sample reproducibility. Intensities were normalized and log-transformed prior to applying Student's $t$-test to determine statistical significance ($p < 0.05$). The $p$-values were adjusted using the Benjamini-Hochberg multiple testing correction. All statistical analyses were performed using the t-test_ind function from the SciPy library in Python (version 3.7.6). KEGG database was utilized for protein annotation of DEPs, and pathway enrichment analysis was performed using a two-sided Fisher's exact test conducted with the fisher_exact function from scipy.stats in Python (version 3.7.6). KEGG pathways with an adjusted $p$-value < 0.05 were considered statistically significant.

***Enzyme-Linked Immunosorbent Assay (ELISA)***: The plasma samples from twin PE cases and healthy controls were thawed and centrifuged at 12,000 rpm for 10 min at 4°C. The supernatants were analyzed using the following ELISA kits: human ANXA1 ELISA kit (ELK Biotechnology, ELK3018), human ANXA2 ELISA kit (ELK Biotechnology, ELK2696), human ANXA3 ELISA kit (ELK Biotechnology, ELK3218), human CHIT1 ELISA kit (ELK Biotechnology, ELK4932), human FTL ELISA kit (ELK Biotechnology, ELK3120), and human S100-A12 ELISA kit (Abcam, ab213822), according to the manufacturers' instructions.

For detection of IL-6 and IL-8 secretion levels, the supernatants from HUVEC cells transfected with siRNA after 48 hours were analyzed using the following ELISA kits: human IL-6 ELISA kit (Ruixinbio, RX106126H) and human IL-8 ELISA kit (Ruixinbio, RX101318H), according to the manufacturers' instructions.

***Cell Culture and Transfection***: HTR8/SVneo cells (ATCC, CRL-3271, RRID: CVCL_7162 HTR8), generously provided by Prof. Yanling Wang from the Institute of Zoology, Institute for Stem Cell and Regeneration, Chinese Academy of Sciences in 2022, were cultured in RPMI 1640 medium (Invitrogen, 22400089). HUVEC cells (kindly provided by Dr. Yang Liu from the Central Laboratory of Peking University Third Hospital in 2022, RRID: CVCL_9Q53) were maintained in DMEM medium (Gibco, C11995500BT). All media were supplemented with 10% fetal bovine serum (Gibco, 10099141C) and penicillin-streptomycin (100 U/mL and 100 μg/mL, HyClone, SV30010). Cells were maintained at 37 °C in a 5% $CO_2$ humidified incubator. Cells were regularly screened for mycoplasma (Yeasen, 40612ES25).

For knockdown of ANXA2, custom-designed siRNAs (siANXA2-1: 5’-CGGCTGTATGACTCCATGA-3’, siANXA2-2: 5’-GTCTGTCAAAGCCTATACT-3’) were purchased from RiboBio (China). When cells reached approximately 80% confluence in six-well plates, each well was transfected with 50 nmol siRNA using Lipofectamine RNAiMAX transfection reagent (#13778150, Thermo Fisher Scientific, USA), which was diluted in Opti-MEM medium (#31985070, Thermo Fisher Scientific, USA). For overexpression of ANXA2, the pCMV plasmid encoding the ANXA2 gene was obtained from Mailgene (Mailgene, MH00460). At approximately 80% confluence in six-well plates, cells in each well were transfected with 3 μg plasmid using transfection reagent Lipofectamine™ 3000 (Invitrogen, L3000015). All the experiments were prepared in triplicate.

***Reverse Transcription Quantitative Real-Time Polymerase Chain Reaction (RT-qPCR)****:* Total RNA was extracted from HTR8/SVneo cells using the RNA Extraction Reagent (Servicebio, G3013). cDNA was synthesized through reverse transcription using the SweScript All-in-One RT SuperMix for qPCR (Servicebio, G3337). RT-qPCR was performed using 2 × SYBR Green Master Mix (Servicebio, G3326) following the manufacturer's instructions. Relative expression levels were calculated using the $2^{-\Delta\Delta Ct}$ quantification method that normalized to *GAPDH* expression. The following primers were used: *GAPDH,* 5’-GGAAGCTTGTCATCAATGGAAATC-3’ and 5’-TGATGACCCTTTTGGCTCCC-3’; *ANXA2,* 5’-ATATTGCCTTCGCCTACCAGAG-3’ and 5’-GCGTCATACTGAGCAGGTGTCTT-3’.

***Western Blotting****:* Total proteins from HTR8/SVneo cells were extracted using RIPA lysis buffer (Servicebio, G2002-100ML) supplemented with PMSF (Servicebio, G2008-1ML), protease inhibitor cocktail (Servicebio, G2006-250UL), and phosphatase inhibitor cocktail (Servicebio, G2007-1ML). Protein concentration was determined using BCA protein assay kit (Servicebio, G2026-200T). Each sample (20 μg of protein) was resolved by SDS-PAGE and subsequently transferred to polyvinylidene fluoride (PVDF) membrane. The membranes were blocked with 5% non-fat milk solution and incubated overnight at 4 °C with anti-ANXA2 antibodies (1:1000, Abcam, ab321903) and anti-GAPDH (1:5000, TransGen Biotech, HC301-02, RRID: AB_2629434) antibodies. After three times washing with TBST, the membranes were further incubated with the secondary anti-rabbit IgG antibodies (1:5000, ZSGB-BIO, ZF-0311, RRID: AB_2571576) or anti-mouse IgG antibodies (1:5000, ZSGB-BIO, ZF-0312, RRID: AB_2716306) at room temperature for 1h. Bands were visualized using ECL reagent (Servicebio, G2014-50ML) and quantitatively analyzed with ImageJ software. The corresponding uncropped gels were presented in Supplemental Figure 9.

***Wound-healing and Transwell Assays***: For HTR8/SVneo cell wound-healing assays, downregulation of ANXA2 by siRNA or upregulation by plasmid transfection were performed. After 24 hours, the transfected cells were seeded into a culture-insert (Ibidi, 81176) at a density of $6 \times 10^4$ cells per well. After 24-hours attachment, the culture-inserts were removed, and the cells were gently washed with PBS. Subsequently, 1 mL of serum-free RPMI 1640 medium was added. Images of the cell-free gap were captured at 0 hour and after 24 hours (for ANXA2 overexpression) or 48 hours (for ANXA2 knockdown) using a light microscope at 50 × magnification.

For transwell assays, 100 μL of serum-free cell suspension ($8\times10^5$ cells/mL) was seeded into the upper chamber (Corning, 3422), and 600 μL medium containing 10% FBS was added to the lower chamber. After incubation for 48 hours (for ANXA2 overexpression) or 72 hours (for ANXA2 knockdown), the invading cells were fixed with 4% paraformaldehyde (PFA; Servicebio, G1101) and stained with 0.1% crystal violet

(BKMAN, 110703008). Images of stained cells were captured under a light microscope at 100 × magnification, and quantitatively analyzed with ImageJ software.

***Immunohistochemistry (IHC) Staining***: The twin placental tissue shares obtained during caesarean deliveries were collected from four normal and four PE twin pregnancies exclusively for immunohistochemistry validation. Population characteristics and pregnancy outcomes were documented (Supplemental Table 3). Placental tissues were extensively washed in PBS, fixed with 4% PFA, and embedded in paraffin. The paraffin-embedded were sectioned by microtome (Leica, RM2016, Shanghai, China) followed by dewaxing with an eco-friendly deparaffinization solution (Servicebio, G1128) and rehydration with gradient alcohol. Antigen retrieval was performed by treatment with 3% hydrogen peroxide and EDTA buffer. After being blocked in goat serum for 20 min, the sections were incubated overnight at 4°C with the following primary antibodies: anti-ANXA2 (1:500, Proteintech, 11256-1-AP, RRID: AB_2057311), anti-ANXA3 (1:500, Proteintech, 11804-1-AP, RRID: AB_2057455), anti-CHIT1 (1:500, Proteintech, 21432-1-AP, RRID: AB_2878857), and anti-FTL (1:100, Abcam, ab313563). After incubation with secondary anti-rabbit IgG antibodies (1:200, Servicebio, GB23303, RRID: AB_2811189) for 30 min at room temperature, the immunoreactive signals were visualized using DAB (3,3'-diaminobenzidine) chromogen solution, followed by washing with distilled water to stop the reaction. Sections were counterstained with hematoxylin, dehydrated in an ethanol gradient, cleared in xylene, and mounted with a neutral resin. Three fields were randomly selected and semi-quantified using ImageJ software.

***Malondialdehyde (MDA) Analyses***: Placental shares from twins were homogenized and lysed in lysis buffer (Beyotime, P0013) with a tissue-to-reagent ratio of 1:9 (g:mL).

The homogenized samples were centrifuged at 10,000 × *g* for 10−15 min at 4°C, and the supernatant was collected for MDA detection following the manufacturer's instructions (Beyotime, S0131S).

***Quantification and Statistical Analyses***: For maternal characteristics, continuous variables were presented as medians with interquartile ranges (IQR), and between-group comparisons were conducted using the Mann–Whitney U test. Categorical variables were summarized as frequencies and percentages, with between-group differences analyzed using the chi-square test or Fisher's exact test. For experimental measurements, continuous variables with a normal distribution (confirmed by Shapiro–Wilk test) were expressed as mean ± standard error of the mean (SEM). Between-group comparisons were performed using Student's t-test for two-group comparisons, one-way analysis of variance (ANOVA) for multi-group comparisons, and two-way ANOVA for comparisons involving both group and time factors. Homogeneity of variance was verified by Levene's test. Post hoc analyses were performed using Tukey's test for equal variances or Dunnett's T3 correction for unequal variances. Statistical analyses were performed using GraphPad Prism version 10.3.1 and R Statistical Software (v4.4.0). A two-sided $P < 0.05$ was considered statistically significant.

***Construction of Prediction Model:*** We employed six distinct machine learning algorithms, including logistic regression, decision tree, naive Bayes, support vector machine (SVM), XGBoost, and k-nearest neighbors (KNN), to evaluate the predictive performance of proteomics-derived biomarkers (CHIT1, KV116, ALB, LTBP2, ANXA1, ANXA3, FTL, and S100-A12) for twin pregnancies with PE. These proteins were selected based on differential expression in the training set (26 PE vs. 34 controls), and all predictive modeling and validation were performed in an independent validation set (14 PE vs. 10 controls) to avoid data leakage. Initially, each biomarker was assessed individually in univariable models. Subsequently, the top five most differentially expressed markers, namely CHIT1, ANXA1, ANXA3, FTL, and S100-A12, were integrated into multivariable models using the same set of machine learning methods. To validate the proteomics findings, we performed ELISA assays to quantify selected protein levels. Based on its potential biological relevance in the pathogenesis of twin PE, ANXA2 was additionally included in the ELISA validation phase. We then constructed models incorporating ANXA2 together with ANXA3, CHIT1, and FTL, again applying all six algorithms. These models were constructed under four configurations: univariable biomarker models (single biomarker), multivariable biomarker models (multiple biomarkers), clinical risk factor models including maternal age, pre-pregnancy BMI, parity (multipara vs. primipara), mode of conception (natural vs. assisted reproductive technology), and mean arterial pressure (MAP = [2 × diastolic pressure + systolic pressure] / 3), and comprehensive models combining biomarkers with these clinical risk factors for PE. Model performance was evaluated using receiver operating characteristic (ROC) curve analysis, with the area under the curve (AUC) and its 95% confidence interval (CI) calculated to assess overall discriminative ability. We report the performance of four model configurations: univariable biomarker models, multivariable biomarker models, clinical risk factor models, and comprehensive models combining biomarkers with clinical risk factors. Sensitivity and specificity for each model were determined at the optimal classification threshold defined by the maximum Youden's Index (J = sensitivity + specificity - 1). The XGBoost algorithm consistently demonstrated superior performance across both proteomic and ELISA datasets. All analyses were conducted using R software (version 4.4.3) with the caret package (version 7.0-1). A two-sided $P < 0.05$ was considered statistically significant.
